# DynaMELD: A Dynamic Model of End-Stage Liver Disease for Equitable Prioritization

**DOI:** 10.1101/2024.11.19.24316852

**Authors:** Michael J. Cooper, Xiang Gao, Xun Zhao, Dariia Khoroshchuk, Yingke Wang, Amirhossein Azhie, Maryam Naghibzadeh, Sandra Holdsworth, Jed Adam Gross, Michael Brudno, Jordan J. Feld, Elmar Jaeckel, Gideon Hirschfield, Rahul G. Krishnan, Mamatha Bhat

## Abstract

Liver transplantation (LT) is a life-saving intervention for patients with end-stage liver disease (ESLD). However, 12–20% of patients listed for LT will die on the waitlist. Modern risk scores used for transplant prioritization cannot encompass the full statistical heterogeneity of patients awaiting LT, disadvantaging women and patients with cholestatic liver disease.

Our study objective was to implement more equitable LT prioritization via a more expressive class of statistical models to individualize risk prediction.

To do so, we created DynaMELD, a deep machine learning-based model of waitlist prioritization. DynaMELD leverages a neural network to model complex interactions between covariates, and leverages the rate-of-change (velocity) of time-varying laboratory biomarkers to predict a more personalized risk of mortality or dropout. Our study cohort comprised 53,046 patients with ESLD listed for LT from 2016– 2023 from the U.S. Scientific Registry of Transplant Recipients.

Using 90-day concordance to measure risk discrimination, DynaMELD achieves 90-day concordance 0.5% higher than MELD 3.0 (*p* < 0.001). Using pooled group concordance (PGCI) as a measure of fairness, DynaMELD achieves a PGCI 1.2% higher for female patients (*p* < 0.001), 8.3% higher for patients with primary biliary cholangitis (*p* < 0.001), 7.2% higher for patients with primary sclerosing cholangitis (*p* < 0.001), and 1.5% higher for patients with acute-on-chronic liver failure Grade 1 (*p* < 0.001) compared to MELD 3.0. DynaMELD reclassifies members of these sub-groups into higher risk tiers, suggesting it would improve their access to organ offers. Introspecting upon DynaMELD using the method of SHapley Additive exPlanations (SHAP) values provides an individualized degree of model interpretability.

Overall, DynaMELD may provide more accurate, individualized predictions of waitlist mortality or dropout to reduce inequities and fairly prioritize patients for liver transplant.

## INTRODUCTION

Liver transplantation (LT) is a life-saving intervention for patients with end-stage liver disease (ESLD). Since organ donation is a limited resource, organ allocation systems attempt to offer organs, as they become available, to the sickest patients occupying the waiting list. This scheme uses a laboratory-derived severity algorithm known as the Model for End-Stage Liver Disease-sodium (MELDNa)^1,2^, which uses sodium, creatinine, bilirubin, and international normalized ratio (INR), and more recently the MELD 3.0, which includes sex and albumin as additional variables, to determine and prioritize the sickest patients for transplant.

By standardizing organ allocation, MELD-based organ allocation has reduced pre-transplant waitlist mortality. Despite this, certain sub-groups of patients, like women^3,4^ and those with primary biliary cholangitis^5^, remain disadvantaged in access to organ offers. These disparities suggest that the MELD model cannot fully encompass the statistical heterogeneity of the patient population listed for liver transplant^6^. Furthermore, previous studies have identified additional covariates with prognostic value, such as serum albumin^7^, patient sex^8^, etiology of liver disease^9^, and frailty^10^, two of which have now been incorporated into the MELD 3.0^11^. Finally, clinicians have hypothesized as early as 2003 that the trajectory of disease progression may also be predictive of pre-transplant mortality, which is not captured by instantaneous MELD^12,13^.

In this study, we hypothesize that key sources of waitlist inequality can be mitigated by employing expressive statistical models that more accurately capture variation in risk among sub-populations. We apply methods from machine learning to create *DynaMELD*, a dynamic and flexible model of end-stage liver disease. DynaMELD employs a neural network to model subtle nonlinear interactions between covariates and risk, and within this framework, we study the incorporation of rates-of-change (the “velocity” and “acceleration”) of laboratory biomarkers to dynamically adjust the score based on the observed trajectory of each individual patient’s disease.

## METHODS

### STUDY DESIGN, POPULATION AND SETTING

Our data were drawn from the Organ Procurement and Transplantation Network (OPTN) Registry, a dataset managed by the Scientific Registry of Transplant Recipients (SRTR) comprising every organ transplant performed in the United States since October 1, 1987. The most recently listed patient in our cohort was registered on March 2, 2023. Use of this dataset was approved by the OPTN, and this study was approved by the Research Ethics Board at University Health Network (REB Study #21-5783).

We structured our cohort to be representative of the patient population prioritized under MELDNa, which was implemented on January 11, 2016. Our cohort comprised adult patients listed for transplant after January 1, 2016, as Hepatitis C represented a dominant indication for transplant in older cohorts^14^. We additionally excluded patients with acute liver failure, patients receiving multi-organ transplant, and patients assigned exception points. Details regarding cohort selection can be found in the Supplement.

Our cohort comprised 53,046 patients. Each patient listed for transplant between January 1, 2016, and December 31, 2018, was assigned into one of the *training*, *validation*, and *test* sub-cohorts, with 80% probability, 10% probability, and 10% probability, respectively. To evaluate the robustness of our method under temporal variation in the data, we constructed a *pre-COVID test* cohort comprising patients listed between January 1, 2019, and December 31, 2019, and a *post-COVID test* cohort comprising patients listed for transplant between January 1, 2020 and March 2, 2023. We separated the *pre-COVID test* set from the *post-COVID test* set to account for potential confounding effects of the COVID-19 pandemic on waitlist dynamics^15^. Table 1 presents the characteristics of our cohort at the time of each patient’s listing.

**Table 1:** Characteristics of our selected cohort at the time of each patient’s listing. These summary statistics are computed prior to imputing missing values. The proportion of patients of each race may sum to more than 100%; in the UNOS registry, some patients are multiply listed across racial categories, perhaps due to those patients identifying with more than one racial or ethnic background. These represent the six most common indications for transplant in the *training* set, listed here in decreasing order of propensity, plus primary sclerosing cholangitis (PSC). To obtain these numbers for PSC, we have grouped together the four diagnosis codes corresponding to PSC: Crohn’s Disease, PSC: Ulcerative Colitis, PSC: No Bowel Disease, and PSC: Other.

|  | Overall<br>(N = 53,046) | Training<br>(N = 15,373) | Validation<br>(N = 1,922) | Test<br>(N = 1,922) | Pre-COVID Test<br>(N = 7,513) | Post-COVID Test<br>(N = 26,316) |
| --- | --- | --- | --- | --- | --- | --- |
| **Demographics** |  |  |  |  |  |  |
| Age, y (IQR) | 54.2 (47.4, 62.8) | 54.8 (48.6, 62.9) | 55.1 (49.4, 63.0) | 54.7 (48.3, 62.8) | 54.9 (48.3, 63.3) | 53.6 (46.0, 62.5) |
| Female, n (%) | 21,503 (40.5%) | 6,197 (40.3%) | 806 (41.9%) | 786 (40.9%) | 3,061 (40.7%) | 10,653 (40.5%) |
| Height, m (IQR) | 1.7 (1.6, 1.8) | 1.7 (1.6, 1.8) | 1.7 (1.6, 1.8) | 1.7 (1.6, 1.8) | 1.7 (1.6, 1.8) | 1.7 (1.6, 1.8) |
| Weight, kg (IQR) | 86.1 (70.6, 98.9) | 86.1 (70.8, 98.9) | 86.6 (70.8, 99.8) | 86.5 (70.8, 99.5) | 86.3 (70.8, 99.8) | 86.0 (70.3, 98.8) |
| **Race<sup>†</sup>** |  |  |  |  |  |  |
| White, n (%) | 39,807 (75.0%) | 11,606 (75.5%) | 1,475 (76.7%) | 1,421 (73.9%) | 5,654 (75.3%) | 19,651 (74.7%) |
| Black, n (%) | 3,234 (6.1%) | 987 (6.4%) | 116 (6.0%) | 122 (6.3%) | 438 (5.8%) | 1,571 (6.0%) |
| Asian, n (%) | 1,591 (3.0%) | 428 (2.8%) | 66 (3.4%) | 67 (3.5%) | 235 (3.1%) | 795 (3.0%) |
| Hispanic/Latino, n (%) | 8,596 (16.2%) | 2,406 (15.7%) | 274 (14.3%) | 316 (16.4%) | 1,204 (16.0%) | 4,396 (16.7%) |
| American Indian, n (%) | 814 (1.5%) | 236 (1.5%) | 24 (1.2%) | 33 (1.7%) | 116 (1.5%) | 405 (1.5%) |
| Native Hawaiian, n (%) | 106 (0.2%) | 27 (0.2%) | 3 (0.2%) | 3 (0.2%) | 17 (0.2%) | 56 (0.2%) |
| **Indication for Transplant<sup>‡</sup>** |  |  |  |  |  |  |
| Alcoholic Cirrhosis, n (%) | 19,478 (36.7%) | 5,634 (36.6%) | 681 (35.4%) | 700 (36.4%) | 2,994 (39.9%) | 9,469 (36.0%) |
| Cirrhosis, MASH, n (%) | 12,070 (22.8%) | 3,478 (22.6%) | 457 (23.8%) | 426 (22.2%) | 1,846 (24.6%) | 5,863 (22.3%) |
| Cirrhosis, Hepatitis C, n (%) | 3,195 (6.0%) | 1,433 (9.3%) | 186 (9.7%) | 194 (10.1%) | 444 (5.9%) | 938 (3.6%) |
| Cirrhosis, Cryptogenic, n (%) | 2,507 (4.7%) | 886 (5.8%) | 120 (6.2%) | 123 (6.4%) | 362 (4.8%) | 1,016 (3.9%) |
| Primary Biliary Cholangitis (PBC), n (%) | 1,754 (3.3%) | 581 (3.8%) | 67 (3.5%) | 75 (3.9%) | 269 (3.6%) | 762 (2.9%) |
| Cirrhosis, Autoimmune, n (%) | 1,845 (3.5%) | 556 (3.6%) | 85 (4.4%) | 80 (4.2%) | 288 (3.8%) | 836 (3.2%) |
| Primary Sclerosing Cholangitis (PSC), n (%) | 2450 (4.6%) | 769 (5.0%) | 82 (4.3%) | 77 (4.0%) | 346 (4.6%) | 1176 (4.5%) |
| **Risk Scores** |  |  |  |  |  |  |
| MELD, mean (IQR) | 21.9 (15.0, 29.0) | 21.9 (15.0, 29.0) | 22.1 (15.0, 29.0) | 22.5 (15.0, 29.0) | 21.6 (14.0, 28.0) | 22.0 (15.0, 29.0) |
| MELDNa, mean (IQR) | 22.9 (15.0, 30.0) | 22.8 (15.0, 30.0) | 22.9 (16.0, 30.0) | 23.4 (16.0, 31.0) | 22.5 (15.0, 30.0) | 23.0 (16.0, 30.0) |
| MELD 3.0, mean (IQR) | 24.0 (16.0, 31.0) | 23.9 (16.0, 31.0) | 24.1 (17.0, 31.0) | 24.6 (17.0, 32.0) | 23.6 (16.0, 30.0) | 24.0 (16.0, 31.0) |
| **Biological** |  |  |  |  |  |  |
| Serum Albumin (g/dL), mean (IQR) | 3.2 (2.7, 3.6) | 3.1 (2.6, 3.6) | 3.1 (2.6, 3.6) | 3.1 (2.6, 3.6) | 3.2 (2.7, 3.6) | 3.2 (2.7, 3.6) |
| Serum Bilirubin (mg/dL), mean (IQR) | 8.9 (2.0, 11.4) | 8.8 (2.0, 10.9) | 8.6 (2.0, 10.5) | 9.4 (2.0, 12.5) | 8.5 (1.9, 10.4) | 9.1 (1.9, 11.8) |
| Serum Creatinine (mg/dL), mean (IQR) | 1.5 (0.8, 1.7) | 1.5 (0.8, 1.7) | 1.5 (0.8, 1.7) | 1.6 (0.8, 1.7) | 1.5 (0.8, 1.7) | 1.5 (0.8, 1.7) |
| Serum Sodium (mmol/dL), mean (IQR) | 135.3 (132.0, 139.0) | 135.5 (132.0, 139.0) | 135.4 (132.0, 139.0) | 135.3 (132.0, 139.0) | 135.3 (132.0, 139.0) | 135.1 (132.0, 139.0) |
| INR, mean (IQR) | 2.0 (1.3, 2.3) | 2.0 (1.3, 2.3) | 2.0 (1.3, 2.3) | 2.0 (1.4, 2.3) | 2.0 (1.3, 2.3) | 2.0 (1.3, 2.3) |
| Ascites (1-4), mean (IQR) | 2.2 (2.0, 3.0) | 2.2 (2.0, 3.0) | 2.2 (2.0, 3.0) | 2.2 (2.0, 3.0) | 2.2 (2.0, 3.0) | 2.2 (2.0, 3.0) |
| Encephalopathy (1-4), mean (IQR) | 1.9 (1.0, 2.0) | 1.9 (1.0, 2.0) | 1.9 (1.0, 2.0) | 1.9 (1.0, 2.0) | 1.9 (1.0, 2.0) | 1.9 (1.0, 2.0) |
| **Outcomes** |  |  |  |  |  |  |
| Pre-Transplant Mortality, n (%) | 4,870 (9.2%) | 1,900 (12.4%) | 239 (12.4%) | 229 (11.9%) | 792 (10.5%) | 1,710 (6.5%) |
| Too Sick to Transplant, n (%) | 3,656 (6.9%) | 1,427 (9.3%) | 189 (9.8%) | 179 (9.3%) | 548 (7.3%) | 1,313 (5.0%) |
| Received Transplant, n (%) | 31,658 (59.7%) | 9,088 (59.1%) | 1,156 (60.1%) | 1,168 (60.8%) | 4,671 (62.2%) | 15,575 (59.2%) |
| Condition Improved, n (%) | 2,965 (5.6%) | 1,243 (8.1%) | 137 (7.1%) | 142 (7.4%) | 506 (6.7%) | 937 (3.6%) |
| Censored (other), n (%) | 9,897 (18.7%) | 1,715 (11.2%) | 201 (10.5%) | 204 (10.6%) | 996 (13.3%) | 6,781 (25.8%) |

### COVARIATE SELECTION AND PREPROCESSING

Our model incorporates serum albumin, creatinine, bilirubin, INR, and sodium into the score. To improve sex-based fairness, our model additionally incorporates patient sex, and to improve diagnosis-based fairness, our model incorporates two indicator variables, reflecting a diagnosis of primary sclerosing cholangitis (PSC), and primary biliary cholangitis (PBC), respectively. This incorporation of diagnosis indicator variables is reminiscent of how the first version of MELD included an indication of whether the patient’s diagnosis was either alcoholic or cholestatic^16^. We imputed missing values using multiple imputation with chained equations^17^. Longitudinal laboratory measurements were incorporated by treating each patient observation as a separate data sample; this of observations is considerably larger than the number of patients.

In keeping with the methodology of the MELDNa and MELD 3.0, we applied a *log*-transformation to right-skewed biological variables (creatinine, bilirubin, INR) before passing them into the model, and applied *z*-score normalization to remaining covariates that were not binary-valued (e.g., rates-of-change). Unlike the MELDNa and MELD 3.0, we did not restrict the values of biomarkers that lay above or below a certain threshold. In the context of models like the MELDNa and MELD 3.0 – *log*-linear models with explicitly defined interaction terms – this thresholding improves model fit by artificially linearizing the covariates. In the context of a nonlinear, neural network-based approach like ours, we did not believe such truncation to be necessary.

Extending prior work that considers the rate-of-change of a patient’s MELD score as a predictive risk factor^12,13^, our approach leverages the rates-of-change of individual biomarkers as a means of learning nuanced relationships between these dynamics and patient risk. To do so, we explore including estimates of the first- and second-order rates-of-change of laboratory-measured values as additional covariates. Figure 1 illustrates how these values were calculated for each laboratory variable (e.g., INR). We calculated the first-order rate-of-change by dividing the difference between prior and current *log*-INR levels (in this case, the *log*-INR at *t*_2_ and *t*_4_) by the amount of time elapsed between observations. This reflects how quickly the patient’s *log*-INR changes as a function of time (e.g., the “velocity” of *log*-INR). We calculated the second-order rate-of-change by dividing the first-order rate-of-change ((*m*_*t*_3__ and (*m*_*t*_4__ in Figure 1) by the amount of time elapsed between observations. This reflects how quickly the rate-of-change of the patient’s *log*-INR changes as a function of time (e.g., the “acceleration” of *log*-INR). At time steps without sufficient prior history to calculate a desired quantity (e.g., calculating the first-order rate-of-change at *t*_1_), we imputed these values with zero. To prevent trends being extrapolated to long periods of time from few samples, we replaced with zero any rate-of-change covariate with a denominator greater than 60 days.

**Figure 1:**
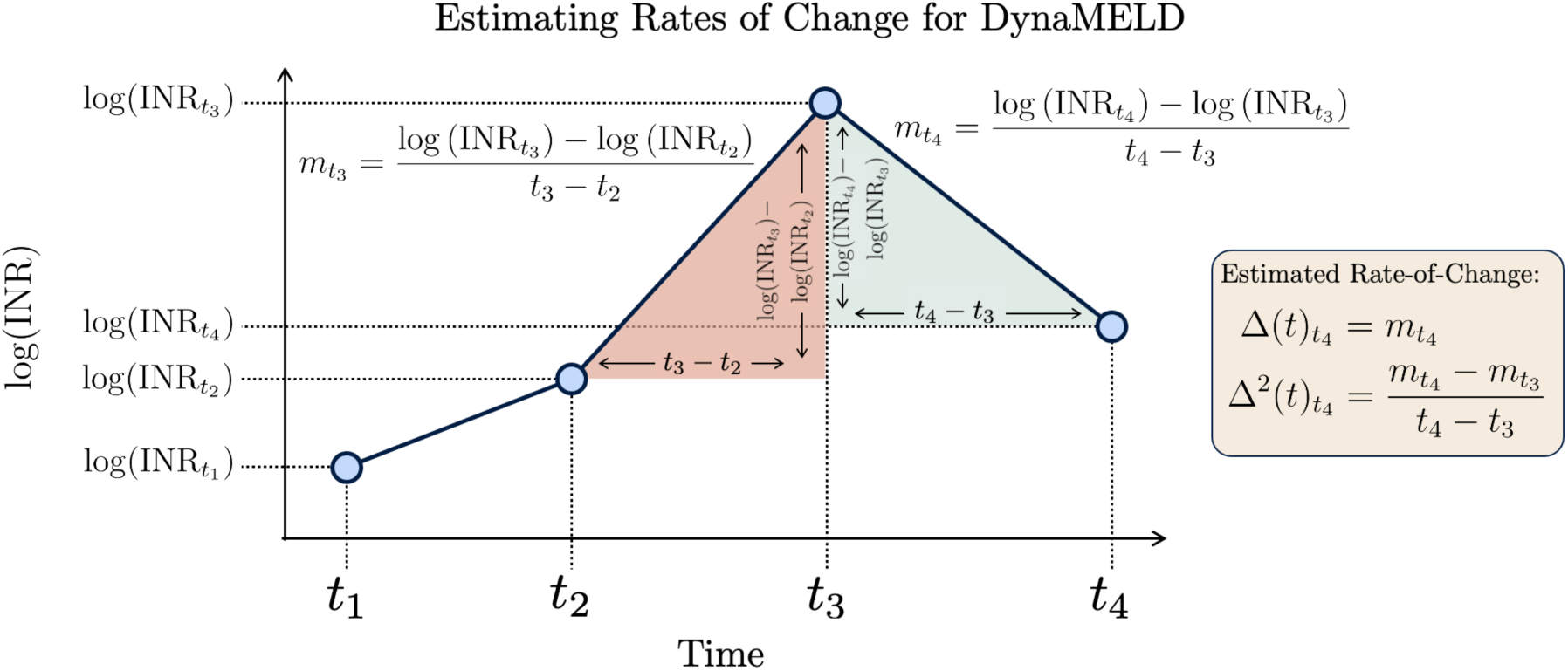
Visualization of how rates-of-change covariates are estimated for use in the DynaMELD model. First-order rates-of-change, such as *m*_*t*_4__, are estimated by dividing the difference in *log*-INR between the current (*t*_4_) and prior (*t*_3_) time steps by the difference in time that has elapsed between them. Second-order rates-of-change are estimated by applying the first procedure recursively to the computed first-order rates-of-change.

### STATISTICAL MODEL

Our outcome of interest was patient time-to-mortality or dropout on the waitlist for being too sick to transplant. Patients who received transplant, withdrew from the waitlist for other reasons, or whose listing surpassed March 2, 2023, are treated as right censored.

DynaMELD is built upon the DeepSurv architecture^18^, a machine learning model that incorporates a neural network into the proportional hazards framework^19^ underlying the MELDNa and MELD 3.0. This approach relaxes the assumption in the Cox model that the *log*-hazard is a linear function of the covariates. Like the Cox model, DeepSurv is trained by maximizing the Cox partial likelihood; however, we additionally regularized the model weights by adding a ridge regularization^20^ term to the loss function to encourage the network weights to take on small absolute values. Empirically, this improves model generalization and training stability. We implemented DynaMELD using the PyTorch Lightning^21^ package, and fit the model to the *training* dataset using the Adam optimizer^22^ with validation-based early stopping^23^ on 90-day concordance^24^. We trained one DynaMELD model for each set of covariates (with and without first- and second-order rate-of-change covariates), and for each set of hyperparameters drawn from a broad hyperparameter search space (e.g., number of hidden layers in the network). After training, we rescaled the score using percentile matching to fit the distribution of the MELD 3.0. Upon observing that patients with PSC remained inadequately ranked with respect to the aggregate population (as measured by pooled group concordance, below), we added four DynaMELD exception points to each observation diagnosed with PSC. The next section reports the results of the DynaMELD model whose hyperparameters achieved the highest 90-day concordance on the *validation* set.

## RESULTS

In this section, we use the *pre-COVID test* set as our primary evaluation set, as we believe it strikes the best balance of highlighting model performance under temporal distribution shift (unlike the *test set*, whose patients were listed at the same time as those in the *training* set), while remaining insulated from confounding effects of the COVID-19 pandemic (unlike the *post-COVID test* set). Results from the *test* set and *post-COVID* test sets can be found in the Supplement. As we are interested in testing noninferiority with respect to the baseline scores, we tested hypotheses using one-tailed *t*-tests comparing the distributions of 100 bootstrap resamples of model performance on the specified evaluation set(s). We set the significance level) to 0.05, and adjusted for multiple comparisons by applying the Holm-Bonferroni correction^25^ to all *p*-values reported in the work.

### RELATIVE RISK DISCRIMINATION

We measure relative risk discrimination using the 90-day concordance index^24^. Of the set of patient pairs for which one or both members experienced mortality/dropout within 90 days, the index summarizes the proportion for whom the higher-risk member is assigned a higher score. This index ranges between 0 and 1, where a higher score indicates a better model: a model that achieves concordance higher than 0.8 is considered to have “excellent diagnostic accuracy” in practical clinical settings^16^.

Figure 2 (Top Left) highlights the 90-day concordance of DynaMELD on the *pre-COVID test* set. On this cohort, DynaMELD (w/ first-order rates-of-change) achieves a 1.2% higher 90-day concordance than the MELDNa (*p* < 0.001) and a 0.5% higher 90-day concordance then the MELD 3.0 (*p* < 0.001). Although the improvement in concordance is modest, it is statistically significant. Incorporating rates-of-change into DynaMELD does not yield statistically significant improvement in 90-day concordance. However, because first-order rates-of-change do not harm discrimination in this setting, and because clinical experience speaks to the importance of these dynamic changes in assessing risk, we report results associated with the *DynaMELD* model that incorporates first-order rate-of-change covariates.

**Figure 2:**
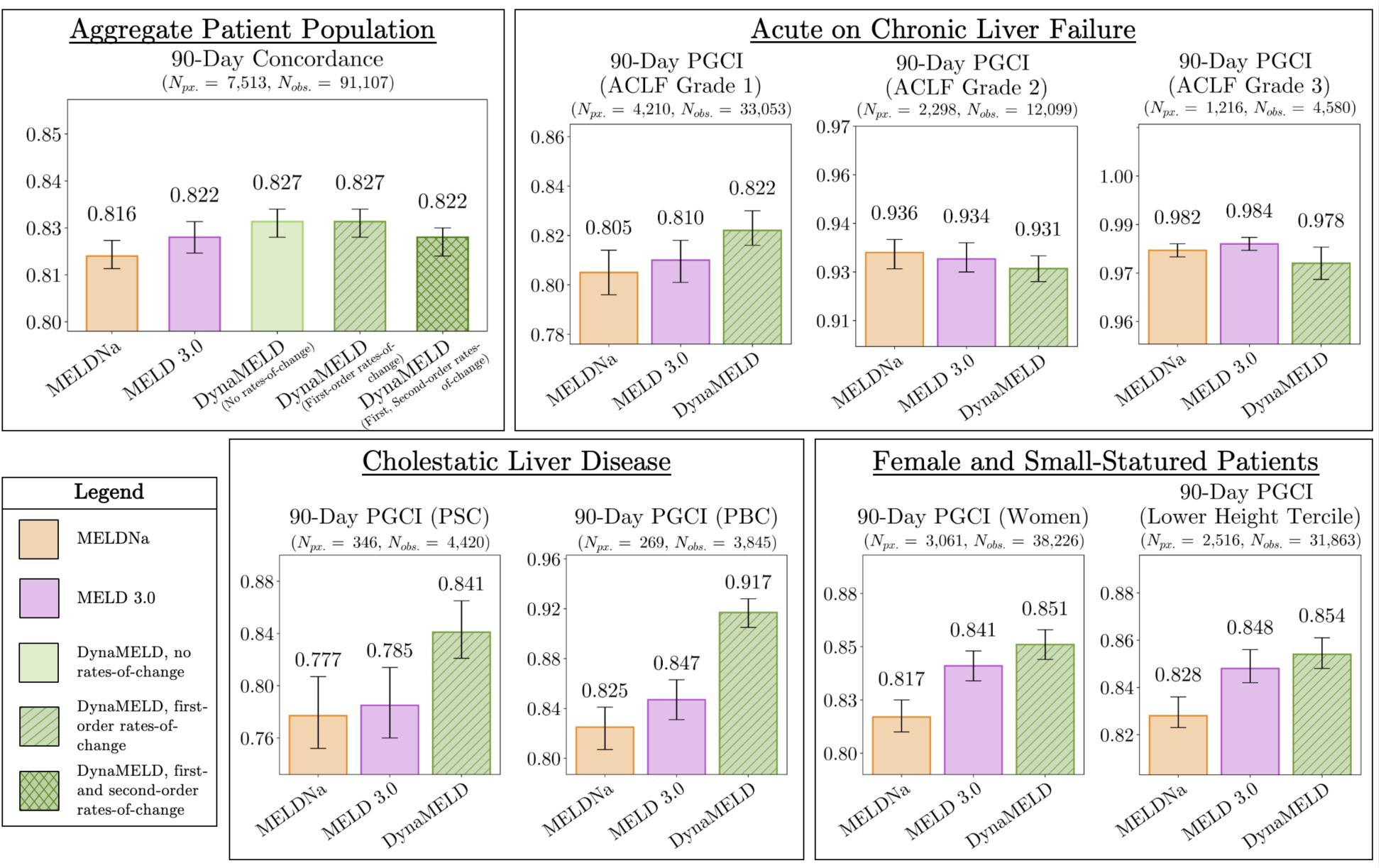
(Top Left) 90-day concordance on the *pre-COVID test* cohort achieved by baselines of the MELDNa, and MELD 3.0, compared to DynaMELD. This plot reports concordance that is computed over all *instances* (N_obs._), of which there is typically more than one per patient (N_px._). The leftmost green bar in this plot represents the DynaMELD model trained with <u>no</u> rate-of-change covariates; the middle bar represents the DynaMELD model trained with <u>first-order</u> rate-of-change covariates; and the rightmost bar represents the DynaMELD model trained with <u>both first- and second-order</u> rate-of-change covariates. DynaMELD achieves statistically significant improvement in 90-day concordance compared to the baselines, although incorporating rate-of-change covariates does not improve risk discrimination in this setting. (Top Right; Bottom) 90-day pooled group concordance (PGCI) achieved by baselines of the MELDNa, and MELD 3.0, compared to DynaMELD. Each plot represents the PGCI for a different identified sub-group on the *pre-COVID test* set: patients with acute-on-chronic liver failure (ACLF; top right), cholestatic liver disease (primary sclerosing cholangitis (PSC) / primary biliary cholangitis (PBC); bottom left), and female / small-statured patients (bottom right). DynaMELD improves PGCI for patients with low-grade ACLF, patients with PSC, patients with PBC, and female and small-statured patients; all of these improvements are statistically significant. The MELDNa and MELD 3.0 achieve modestly higher PGCI in patients with ACLF Grades 2 and 3.

### FAIRNESS AND RECLASSIFICATION

The 90-day pooled group concordance index (PGCI)^26^ provides a statistical means to assess the fairness of survival models. It modifies 90-day concordance to restrict comparison to those pairs whose higher-risk member belongs to an *identified sub-group* (e.g., women). Loosely, the PGCI assesses the model’s ability to correctly rank members of the identified sub-group by risk against the rest of the patient population. Figure 2 (Bottom) shows that DynaMELD achieves a higher PGCI than MELDNa and MELD 3.0 in patients with primary sclerosing cholangitis (PSC; *p* < 0.001), patients with primary biliary cholangitis (PBC; *p* < 0.001), female patients (*p* < 0.001), and small-statured patients (patients shorter than 166 cm (5’.5”), the lower tercile of the training set; *p* < 0.001).

We additionally analyzed how DynaMELD would have reclassified decedents on the waitlist. We calculated the risk percentile associated with the scores assigned by DynaMELD and the MELD 3.0. Then, we selected the subset of the cohort who did not receive an organ and examined their assigned risk quantiles. A score that places this cohort of patients into a higher risk percentile is desirable, as it may have afforded these decedents – who were under-served by the MELDNa – a higher chance of receiving transplant. This analysis was repeated for women, patients with PSC, patients with PBC, and small-statured patients. These results are shown in Figure 3: observe how DynaMELD places more members of these sub-groups into higher risk quantiles, suggesting that it would have afforded these patients better access to organ offers.

**Figure 3:**
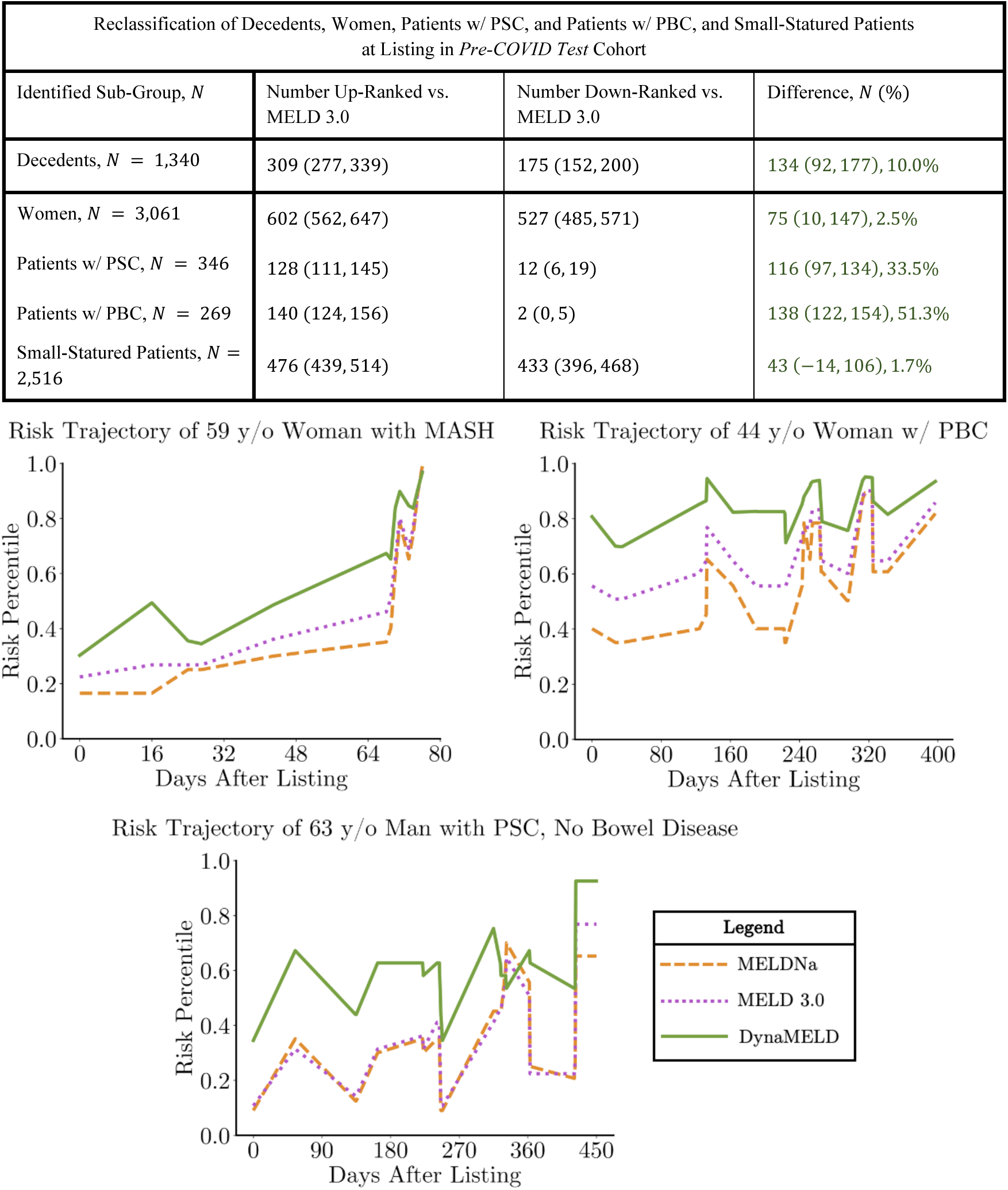
(Top) In a population of decedents from the *pre-COVID test* cohort (prioritized under MELDNa), we study the reclassification of patients into different risk quintiles under the MELD 3.0 and DynaMELD. Relative to the MELD 3.0, DynaMELD assigns 309 decedents a higher risk quintile, compared to 175 who are placed into a lower quintile. Under DynaMELD, 10.0% more of these decedents would have been afforded a higher chance of receiving a donor organ prior to their time of death. Parentheses represent 95% confidence intervals computed from 1,000 bootstrap resamples. A similar reclassification argument can be made for members of disadvantaged sub-groups. DynaMELD classifies 2.5% more women, 33.5% more PSC patients, 51.3% more PBC patients, and 1.7% more small-statured patients in the *pre-COVID test* set into a higher risk quintile than the MELD 3.0. (Bottom) Selected individual risk trajectories of patients under DynaMELD, MELDNa and MELD 3.0. All three patients died on the waitlist, after 77 days, 498 days, and 397 days, respectively. Observe how DynaMELD placed these patients into a higher risk percentile earlier on in their trajectory compared to the MELDNa and MELD 3.0, better identifying their risk of deterioration and improving their chance of attracting a donor organ in time.

### PRIORITIZING PATIENTS WITH ACUTE-ON-CHRONIC LIVER FAILURE

We next investigated the extent to which DynaMELD prioritized patients with acute on chronic liver failure (ACLF)^27^. Because the SRTR registry does not provide the necessary variables to directly apply the EASL criteria^28^, we employed a close proxy to grade instances of ACLF (see the Supplement for our procedure). Applying these criteria found that 54.5% of instances in the *pre-COVID test* set exhibited some degree of ACLF. DynaMELD obtains statistically significant improvements in PGCI on patients with low-grade ACLF, with a PGCI 1.5% higher than the MELD 3.0 in patients with ACLF Grade 1 (*p* < 0.001), and comparable performance to MELD 3.0 in patients with ACLF Grades 2 and 3.

### INTERPRETING MODEL PREDICTIONS

Unlike the coefficients of linear models, the model weights of neural networks cannot be directly interpreted to determine the significance of a covariate to the prediction. Moreover, the significance of each covariate to the prediction is subject to vary between patients. Therefore, we employed the established method of SHAP (SHapley Additive exPlanations) values to interpret the relevance of each covariate to each prediction^29^. The SHAP procedure was calibrated on 5,000 sampled observations from the training set and evaluated using 5,000 sampled observations from the *pre-COVID test* set. Figure 4 (top) shows that DynaMELD places the greatest significance on features included in MELD 3.0, suggesting that much of the improvement in DynaMELD’s performance comes from learning interactions that are not captured by the functional form of MELD 3.0. Moreover, rate-of-change covariates are associated with outlier SHAP values, suggesting that, for some patients, the incorporation of rate-of-change covariates plays an important role in accurate risk prediction. Subsequent analysis reveals that, of the instances in the *pre-COVID test* set with at least one non-negligible SHAP value (abs. val. > 0.2) associated with one or more rate-of-change covariates, 95.1% underwent a score update within the last three days. This suggests that DynaMELD learns that the more significant rates-of-change belong to the sicker (perhaps hospitalized) patients who receive more frequent score updates, and whose rates-of-change contain smaller denominators^30^. Models like MELDNa or MELD 3.0 that rely solely on a patient’s biomarkers at a single point in time would be unable to extract predictive signal of this kind. Figure 4 (bottom) highlights how these SHAP values can be rescaled so that they are denominated in units of (Dyna)MELD points, to provide an individualized interpretation of model predictions.

**Figure 4:**
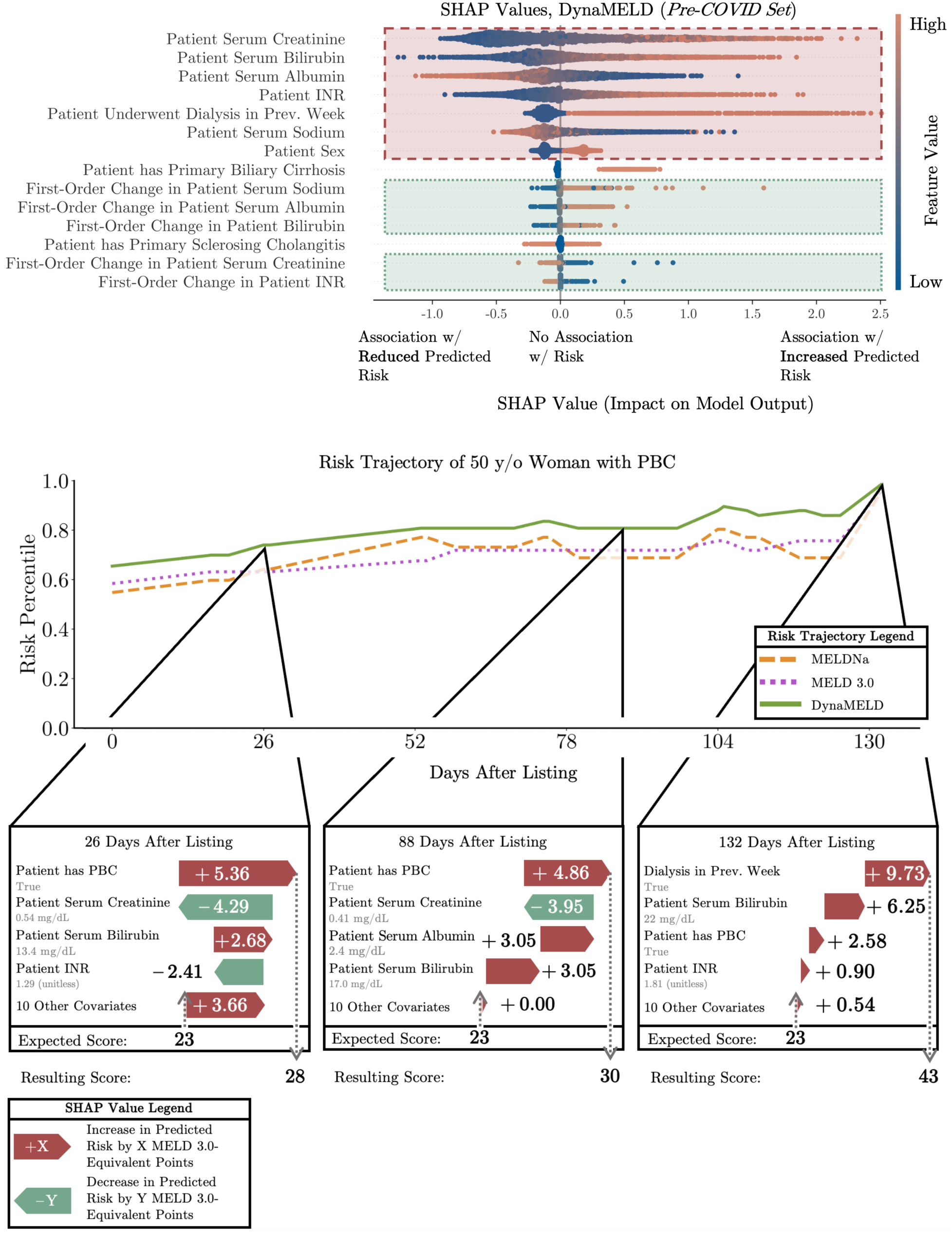
(Top) Swarm plot highlighting the most influential predictive features under DynaMELD, listed in order of decreasing average influence. Each dot represents a single patient observation. The location of each dot on the horizontal axis represents its SHAP value, or the significance of the corresponding feature to the prediction made for that individual patient. The colour of each dot reflects the feature value (e.g., the patient’s serum sodium is low). This plot illustrates that, overall, DynaMELD finds larger values of creatinine, bilirubin, and INR to be correlated with increased risk predictions (because orange dots are typically found farther to the right), while the opposite is true for serum albumin and serum sodium. This same direction of association is found in the learned coefficients of the MELDNa and MELD 3.0, showing that DynaMELD identifies statistical relationships already known to the clinical community. The plot is overlaid with two sets of rectangles. Red dashed rectangles highlight the covariates that comprise the MELD 3.0, including covariates like dialysis status which is implicitly incorporated by reassigning the value of serum creatinine. That these features lie near the top of the graph indicates that these features are of greatest significance in DynaMELD’s predictions. Green dotted rectangles highlight the significance of rate-of-change covariates: although these have low average significance, the presence of clear outliers (e.g., dots far to the left and far to the right) highlights that DynaMELD finds the rate-of-change to provide important predictive signal in some instances. (Bottom) SHAP values provide an individualized degree of model interpretability. The top line chart showcases a selected patient’s risk trajectory under MELDNa, MELD 3.0, and DynaMELD. At three time points of observation, we plot the SHAP values associated with the prediction at that time. The corresponding waterfall plots visualize how an initial population “expected” (average; here, 23) score is up- and down-adjusted based on the contribution of each covariate to arrive at a “resulting score,” representing DynaMELD’s prediction on that instance. Although there is similarity in the most significant predictive factors for this patient over time, there are also key differences: dialysis status, for example, becomes a very significant predictive factor later in this patient’s trajectory, while their diagnosis of PBC is a more significant predictor earlier in their trajectory. This is a degree of statistical nuance that would go largely uncaptured by inspecting the coefficients of a *log*-linear model with explicitly defined interaction terms, like the MELD 3.0.

## DISCUSSION

Over the last twenty years, neither the MELD nor its successors have progressed far beyond linear survival analysis on a limited covariate set to estimate the severity of ESLD. This approach struggles to capture the rich diversity of a heterogenous patient population, and often disadvantages members of certain sub-groups. Moreover, this functional form struggles to harness the complex evolutionary dynamics of time-varying biomarkers, which clinical experience tells us are valuable for accurate risk assessment.

DynaMELD takes a step toward implementing a more accurate, fair system of waitlist prioritization. The use of a nonlinear neural network allows the model to capture subtle interactions between covariates, resulting in a statistically significant improvement to 90-day concordance and PGCI over the MELD 3.0. This suggests that DynaMELD supports more accurate risk-based prioritization and may help mitigate inequities in waitlist outcomes for patients with cholestatic liver disease, female patients, and small-statured patients. Furthermore, by considering the inclusion of several covariates beyond those of the MELD 3.0, such as sparse indicators of primary diagnosis, DynaMELD provides predictions that are more personalized to each patient’s set of characteristics and longitudinal biomarker changes. That the covariates underlying DynaMELD are routinely collected in waitlisted patients across the United States suggests that no changes to established data collection procedures would impede clinical implementation or further study. Finally, DynaMELD is a reliable, interpretable score: analysis of DynaMELD’s SHAP values permits ready interpretation of its predictions for each individual patient, and validates that DynaMELD learns causal relationships that are known to the clinical community.

Our work is not without limitations. First, our cohort excludes patients with exception points, such as those with hepatocellular carcinoma. This was a deliberate decision based on the rationale that exception points represent a parallel prioritization system disjoint from the MELD-based scheme^31,32^. Second, the first-order rates-of-change within DynaMELD introduce a degree of subjective modifiability into the score, because the denominator of each rate-of-change is determined by the amount of time between score updates (Figure 1). It is therefore conceivable that frequently updating a patient’s score may exaggerate the rates-of-change associated with their biomarkers and artificially inflate his or her predicted risk under our model. We recommend that any implementation of DynaMELD be paired with a carefully designed update schedule to reduce subjectivity associated with the timing of score updates. Third, the OPTN registry from which we draw our training data does not require that laboratory MELD scores be immediately sent to the OPTN; permissible delays range from two to thirty days, depending on the MELD score being reported^30^. This means that the times-to-event associated with intermediary MELD updates may exhibit some degree of response (label) error, although this limitation would similarly afflict any time-to-event model that utilizes these intermediary updates. Finally, although DynaMELD improves PGCI in almost all diagnostic and demographic sub-groups studied (see both the main results and Supplement), these improvements are not universal: for example, the MELDNa and MELD 3.0 achieve a higher PGCI on patients with Hepatitis B cirrhosis (*p* < 0.001) – which is less important in the current era – and on patients with higher-grade ACLF. We recommend further investigation of whether assigning exception points to sub-groups beyond those with PSC may mitigate potential lingering inequities under this version of DynaMELD.

Our work opens promising avenues for future scholarship. Future work may study whether a single model can leverage the flexible functional form of DynaMELD to reconcile the disparate prioritization scheme introduced by exception points. Additionally, because the PGCI is a measure of *demand-side inequity* – being insensitive to supply-side factors that may influence fairness – future work may leverage tools like organ allocation simulation to study and mitigate supply-side sources of inequity.

As clinical care becomes increasingly multimodal – relying on combinations of laboratory data, demographic data, and imaging data to support decision-making – there emerges a need to build statistical models of risk that accommodate a diverse combination of heterogenous data modalities. DynaMELD’s basis in neural networks makes it a flexible, forward-looking framework that is well-positioned to incorporate such additional data should the need arise. For example, a modality like radiological images could be readily incorporated into DynaMELD by training a sub-neural network on a dataset of radiological images, then folding the sub-network into DynaMELD via a straightforward “network fusion” operation^33^. In this way, DynaMELD not only accommodates the contemporary needs of the clinical community – by improving predictive accuracy and outcome equity – but establishes a solid foundation for future data-driven decision-making in pre-transplant care.

## Data Availability

The code required to reproduce the results of this manuscript is linked in the article. The data underlying the present study can be requested from the Scientific Registry of Transplant Recipients. (https://www.srtr.org/about-the-data/the-srtr-database/)

