## Supplementary Materials for "DynaMELD: A Dynamic Model of End-Stage Liver Disease for Equitable Prioritization"

(Supplementary Material)

Michael J. Cooper, M.S.[1,2,3], Xiang Gao[1,3], Xun Zhao, M.D.[4], Dariia Khoroshchuk, M.S.[2,3], Yingke Wang B.Sc.[5], Amirhossein Azhie, M.D.[6], Maryam Naghibzadeh, M.D.[7], Sandra Holdsworth[8], Jed Adam Gross, J.D.[9], Michael Brudno, Ph.D.[1,2,10], Jordan J. Feld, M.D.[11], Elmar Jaeckel, M.D.[12,13], Gideon Hirschfield, M.D.[11], Rahul G. Krishnan, Ph.D.[*,1,3,14], Mamatha Bhat, M.D.[*,10,12,13]

Affiliations:

1. Department of Computer Science, University of Toronto, Toronto, ON, Canada
2. University Health Network, Toronto, ON, Canada
3. Vector Institute for Artificial Intelligence, Toronto, ON, Canada
4. Department of Gastroenterology and Hepatology, McGill University, Montreal, QC, Canada
5. Department of Computer Science, University of Waterloo, Waterloo, ON, Canada
6. Max Rady College of Medicine, University of Manitoba, Winnipeg, MB, Canada
7. Kingston Health Sciences Centre, Kingston, ON, Canada
8. Canadian Donation and Transplantation Research Program, Edmonton, AB, Canada
9. Department of Clinical and Organizational Ethics, University Health Network, Toronto, ON, Canada
10. Transplant AI Initiative, Ajmera Transplant Program, Toronto, ON, Canada
11. Toronto Centre for Liver Disease, University Health Network, Toronto, ON, Canada
12. Ajmera Transplant Program, University Health Network, Toronto, ON, Canada
13. Division of Gastroenterology, Department of Medicine, University of Toronto, Toronto, ON, Canada
14. Department of Laboratory Medicine and Pathobiology, University of Toronto, Toronto, ON, Canada

Notes:

[*] Co-senior authors

#### CONTENTS (SUPPLEMENTARY MATERIALS)

#### Appendix A: Code and Model Release

The software code to reproduce the results of this paper, and instructions on obtaining copies of our trained models for further study, can be found here: https://github.com/rgklab/DynaMELD.

#### Appendix B: Cohort Selection

##### Appendix B.1: Cohort Selection and Exclusion Criteria

Our UNOS cohort was defined using the criteria specified in Table S.1. and Table S.2.

| Exclusion Criterion | Number Excluded | Number Remaining |
|---|---|---|
| *Initial Cohort* | – | 355,094 |
| Remove patients listed outside time of interest (prior to January 1, 2016). | 259,013 | 96,081 |
| Remove patients who received transplants but were never put on the waitlist, or who were listed in error. | 0 | 96,081 |
| Remove patients with one or more previous transplants. | 4,575 | 91,506 |
| Remove patients who received multi-organ transplants. | 5,088 | 86,418 |
| Remove patients with acute liver failure. | 3,106 | 83,312 |
| If transplanted, remove patients with HIV. | 317 | 82,995 |
| Remove patients with HCC related primary or secondary diagnosis. | 17,942 | 65,053 |
| Remove patients approved for listing under HCC exception. | 638 | 64,415 |
| Remove patients with exception points for any other reason. | 7,048 | 57,367 |
| Remove patients who are multi-listed, refused transplant, transferred, unable to be contacted, transplanted in another country, or removed in error. | 2,717 | 54,650 |
| Remove non-adult patients (under 18 years of age) and Status 1B patients. | 1,319 | 53,331 |
| Remove patients with pre-listing malignancy if hepatocellular carcinoma, hepatoblastoma, cholangiocarcinoma. | 285 | 53,046 |
| Total Included Cohort | – | 53,046 |

**Table S.1:** Table of exclusion criteria, as well as the number of patients who are excluded as the criteria are sequentially applied to the SRTR dataset.

| Exclusion Criterion | Number Excluded | Number Remaining |
|---|---|---|
| *Initial Cohort* | – | 355,094 |
| Remove patients listed outside time of interest (prior to January 1, 2016).<br>Criterion:<br>CAN_ACTIVATE_DT ≤ January 1, 2016. | 259,013 | 96,081 |
| Remove patients who received transplants but were never put on the waitlist, or who were listed in error.<br>Criterion:<br>CAN_SOURCE = “L” (live donor never on waitlist), *or*<br>CAN_REM_CD = 10 (candidate listed in error). | 0 | 96,081 |
| Remove patients with one or more previous transplants.<br>Criterion:<br>CAN_PREV_TX ≠ 0 (candidate underwent previous transplant), *or*<br>CAN_PREV_HL ≠ 0 (candidate underwent previous heart-lung transplant), *or*<br>CAN_PREV_HR ≠ 0 (candidate underwent previous heart transplant), *or*<br>CAN_PREV_IN ≠ 0 (candidate underwent previous intestine transplant), *or*<br>CAN_PREV_KI ≠ 0 (candidate underwent previous kidney transplant), *or*<br>CAN_PREV_KP ≠ 0 (candidate underwent previous kidney-pancreas transplant), *or*<br>CAN_PREV_LI ≠ 0 (candidate underwent previous liver transplant), *or*<br>CAN_PREV_LU ≠ 0 (candidate underwent previous lung transplant), *or*<br>CAN_PREV_PA ≠ 0 (candidate underwent previous pancreas transplant). | 4,575 | 91,506 |
| Remove patients who received multi-organ transplants.<br>Criterion:<br>REC_TX_ORG_TY ≠ “LI” (candidate received any organ other than only a liver). | 5,088 | 86,418 |
| Remove patients with acute liver failure (Status 1A).<br>Criterion:<br>CANHX_STAT_CD = 1010 (Status 1A), *or*<br>CANHX_STAT_CD = 1090 (Old Status 1A), *or*<br>CANHX_STAT_CD = 1110 (Adult Status 1A), *or*<br>CANHX_STAT_CD = 2010 (Status 1A), *or*<br>CANHX_STAT_CD = 2110 (Adult Status 1A), *or*<br>CANHX_STAT_CD = 3010 (Status 1), *or*<br>CANHX_STAT_CD = 6010 (Status 1), *or*<br>CANHX_STAT_CD = 6011 (Status 1A), *or*<br>CANHX_STAT_CD = 9010 (Status 1A). | 3,106 | 83,312 |
| If transplanted, remove patients with HIV.<br>Criterion:<br>REC_HIV_STAT = “P” (patient is HIV-positive). | 317 | 82,995 |
| Remove patients with HCC related primary or secondary diagnosis.<br>Criterion:<br>CAN_DGN = 4400 (primary diagnosis code 4400), *or*<br>CAN_DGN2 = 4400 (secondary diagnosis code 4400), *or*<br>CAN_DGN = 4401 (primary diagnosis code 4401), *or*<br>CAN_DGN2 = 4401 (secondary diagnosis code 4401), *or*<br>CAN_DGN = 4402 (primary diagnosis code 4402), *or*<br>CAN_DGN2 = 4402 (secondary diagnosis code 4402), *or*<br>CAN_DGN = 4403 (primary diagnosis code 4403), *or* | 17,942 | 65,053 |

| | | |
|---|---|---|
| CAN_DGN2 = 4403 (secondary diagnosis code 4403), *or*<br>CAN_DGN = 4404 (primary diagnosis code 4404), *or*<br>CAN_DGN2 = 4404 (secondary diagnosis code 4404), *or*<br>CAN_DGN = 4405 (primary diagnosis code 4405), *or*<br>CAN_DGN2 = 4405 (secondary diagnosis code 4405), *or*<br>CAN_DGN = 4410 (primary diagnosis code 4410), *or*<br>CAN_DGN2 = 4410 (secondary diagnosis code 4410), *or*<br>CAN_DGN = 4420 (primary diagnosis code 4420), *or*<br>CAN_DGN2 = 4420 (secondary diagnosis code 4420), *or*<br>CAN_DGN = 4430 (primary diagnosis code 4430), *or*<br>CAN_DGN2 = 4430 (secondary diagnosis code 4430).<br><br>Reference:<br>4400 (diagnosis code) – Hepatoma, Hepatocellular Carcinoma<br>4401 (diagnosis code) – Hepatoma (HCC) and Cirrhosis<br>4402 (diagnosis code) – Fibrolamellar (FL-HC)<br>4403 (diagnosis code) – Cholangiocarcinoma (CH-CA)<br>4404 (diagnosis code) – Hepatoblastoma (HBL)<br>4405 (diagnosis code) – Hemangioendothelioma, Hemangiosarcoma, Angiosarcoma<br>4410 (diagnosis code) – Primary Hepatic Malignancy, Other Specify (i.e., Klatzkin Tumor, Leiomysarcoma)<br>4420 (diagnosis code) – Bile Duct Cancer (Cholangioma, Biliary Tract Carcinoma)<br>4430 (diagnosis code) – Secondary Hepatic Malignancy, Other Specify | | |
| Remove patients approved for listing under HCC exception.<br>Criterion:<br>[CAN_CANHX_MPXCPT_DGN = 1 (MELD exception diagnosis is "HCC Meeting Criteria" (Stage T2)), *or*<br>CAN_CANHX_MPXCPT_DGN = 10 (MELD exception diagnosis is "HCC Meeting Criteria" (Stage T1)), *and*<br>[CAN_CANHX_MPXCPT_STAT = 5 (MELD exception status approved), *or*<br>CAN_CANHX_MPXCPT_STAT = 14 (MELD exception status approved; not extended), *or*<br>CAN_CANHX_MPXCPT_STAT = 16 (MELD exception status approved; newer case), *or*<br>CAN_CANHX_MPXCPT_STAT = 18 (MELD exception status approved; overwritten by higher MELD/PELD), *or*<br>CAN_CANHX_MPXCPT_STAT = 30 (MELD exception status approved; extended)]. | 638 | 64,415 |
| Remove patients with exception points for any other reason.<br>Criterion:<br>CAN_CANHX_MPXCPT_STAT = 5 (MELD exception status approved), *or*<br>CAN_CANHX_MPXCPT_STAT = 14 (MELD exception status approved; not extended), *or*<br>CAN_CANHX_MPXCPT_STAT = 16 (MELD exception status approved; newer case), *or*<br>CAN_CANHX_MPXCPT_STAT = 18 (MELD exception status approved; overwritten by higher MELD/PELD), *or*<br>CAN_CANHX_MPXCPT_STAT= 30 (MELD exception status approved; extended). | 7,048 | 57,367 |
| Remove patients who are multi-listed, refused transplant, transferred, unable to be contacted, transplanted in another country, or removed in error.<br>Criterion:<br>CAN_REM_CD = 14 (Transplant at Another Centre; Multi-Listed), *or*<br>CAN_REM_CD = 6 (Candidate Refused Transplant), *or*<br>CAN_REM_CD = 7 (Candidate Transferred to Another Centre), *or*<br>CAN_REM_CD = 24 (Unable to Contact Candidate), *or*<br>CAN_REM_CD = 22 (Transplanted in Another Country), *or*<br>CAN_REM_CD = 16 (Candidate Removed in Error). | 2,717 | 54,650 |
| Remove non-adult patients (under 18 years of age) and Status 1B patients.<br>Criterion: | 1,319 | 53,331 |

| | | |
|---|---|---|
| CAN_AGE_IN_MONTHS_AT_LISTING < (18 × 12), *or*<br>CANHX_STAT_CD = 1020 (Status 1B), *or*<br>CANHX_STAT_CD = 2020 (Status 1B), *or*<br>CANHX_STAT_CD = 6012 (Status 1B). | | |
| Remove patients with pre-listing malignancy if hepatocellular carcinoma, hepatoblastoma, cholangiocarcinoma.<br>Criterion:<br>CAN_MALIG_TY = 4096 (previous malignancy: hepatocellular carcinoma), *or*<br>CAN_MALIG_TY = 8192 (previous malignancy: hepatoblastoma), *or*<br>CAN_MALIG_TY = 4096 (previous malignancy: cholangiocarcinoma). | 285 | 53,046 |
| Total Included Cohort | – | 53,046 |

**Table S.2:** Table of exclusion criteria, annotated with SAF (Standard Analysis File) Data Dictionary codes for reproducibility. The codes listed in this table indicate the criteria necessary to *exclude* the corresponding members of the cohort: for example, writing "CAN_DGN = 4400" means that we exclude candidates with a primary diagnosis code (CAN_DGN) of 4400 (hepatocellular carcinoma).

##### Appendix B.2: Data Splitting and Composition

Our method treats each patient observation as an independent data sample[†] to learn predictions of time-to-event that apply at any point during a patient's waitlist tenure. Because there is often more than one observation per patient, the number of entries in each data table is substantially larger than the number of patients in each table. The table below summarizes the number of patients and entries in each subset of the data.

[†] Although independence may be violated in the data in instances which are explicitly noted – such as when the rate-of-change of time-varying biomarkers is estimated from prior entries in the data corresponding to the same patient – our methodology makes the simplifying assumption of treating each patient observation as an independent sample.

| Data Table | Number of Patients | Number of Entries (Observations) |
|---|---|---|
| *Training* | 15,373 | 212,873 |
| *Validation* | 1,922 | 27,760 |
| *Test* | 1,922 | 28,074 |
| *Pre-COVID Test* | 7,513 | 91,107 |
| *Post-COVID Test* | 26,316 | 220,789 |
| *Total* | 53,046 | 580,603 |

**Table S.3:** The number of patients and number of entries associated with each split of the SRTR dataset. Observe that there are substantially more entries than patients: in the UNOS cohort (across all splits of the data), each patient is associated with an average of 10.9 entries.

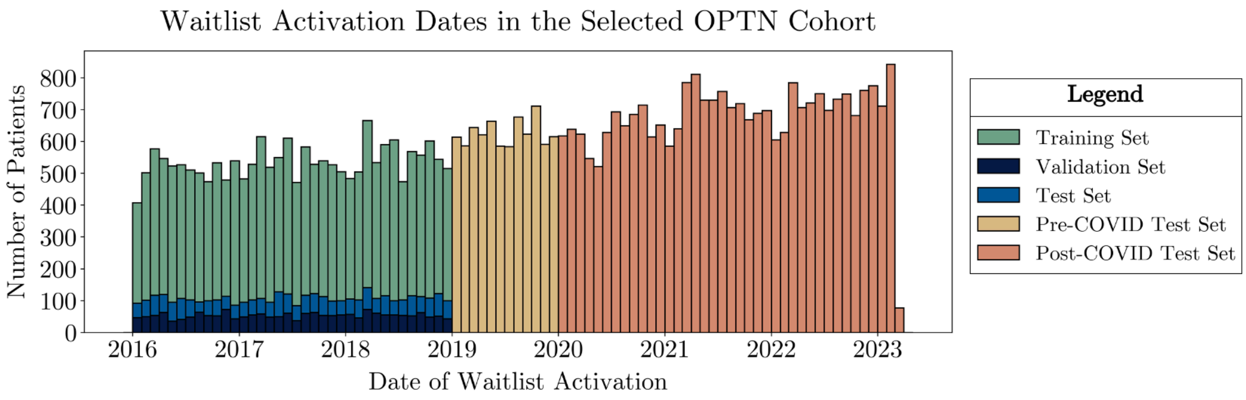

**Figure S.1:** Plot of the listing times of each patient in the selected OPTN cohort. The height of each bar represents the number of patients listed in each month, while the colour of each indicates the subset to which each patient is assigned during data splitting.

#### Appendix C: Selected Features and Preprocessing

**Appendix C.1: Taxonomy of Covariates**

In the following sections, we present the covariates associated with each feature set, along with our processing of each covariate prior to passing into the model. Our covariate processing follows a simple rule-based scheme that depends on the *encoding* and *temporal classification* of each covariate. As such, here we highlight the possible set of encodings and temporal classifications that each covariate can take and explain how the encoding and temporal classification of each covariate influences its preprocessing. Then, in the tables in Appendices C.2. through C.4., we annotate each covariate by its encoding and temporal classification, which will dictate how it is processed.

We encode each covariate in one of the following two ways:

1. Binary: binary variables take on values of either 1 or 0. These represent quantities that are either binary-valued (true or false), or instances in which the patient may fall into one of two discrete categories (e.g., patient sex). We represent such variables using a single column in the data table, where a value of 1 indicates instances in which the quantity is true, and 0 indicates instances where the quantity is false.

2. Continuous: continuous variables represent quantities that can take on a continuous range of values. Such variables are represented by a single column in the data table, where each row represents the value of the variable for that instance.

Each covariate in the data also has one of two temporal classifications:

1. Static: static variables are those assigned to the patient at listing that are assumed not to change over the course of a patient's waitlist tenure. In our case, this means that these variables will remain the same in all entries associated with the same patient in our dataset.

2. Time-Varying: time-varying variables are those that are updated at each patient observation.

3. Time-Varying (Δ): time-varying variables are those that are updated at each patient observation. The (Δ) indicates that these are the variables for which we calculate corresponding first- and second-order rate-of-change covariates during feature selection.

In the tables below, we annotate each feature name with its corresponding SAF (SRTR Standard Analysis File) Data Dictionary code(s) for reproducibility and to reduce ambiguity. In general, codes with the prefix "CAN_" correspond to static covariates drawn from the CAND_LIIN (Candidate: Liver/Intestine) table, whereas codes with the prefix "CANHX_" correspond to time-varying covariates drawn from the STATHIST_LIIN (Status History: Liver/Intestine) table.

#### Appendix C.2: The DynaMELD Covariate Set

| Covariates in the *MELD3.0* Set | | | |
|---|---|---|---|
| Feature Name | Encoding | Temporal Classification | Units |
| Sex<br>SAF Data Dictionary Code(s): CAN_GENDER | Binary<br>1 indicates Female | Static | – |
| Serum Bilirubin<br>SAF Data Dictionary Code(s): CANHX_BILI | Continuous | Time-Varying (Δ) | mg/dL |
| International Normalized Ratio<br>SAF Data Dictionary Code(s): CANHX_INR | Continuous | Time-Varying (Δ) | – |
| Serum Creatinine<br>SAF Data Dictionary Code(s): CANHX_SERUM_CREAT | Continuous | Time-Varying (Δ) | mg/dL |
| Serum Sodium<br>SAF Data Dictionary Code(s): CANHX_SERUM_SODIUM | Continuous | Time-Varying (Δ) | mmol/dL |
| Serum Albumin<br>SAF Data Dictionary Code(s): CANHX_ALBUMIN | Continuous | Time-Varying (Δ) | g/dL |
| Candidate Diagnosis of Primary Sclerosing Cholangitis<br>SAF Data Dictionary Code(s):<br>CAN_DGN = 4240 (primary diagnosis code 4240), *or*<br>CAN_DGN = 4241 (primary diagnosis code 4241), *or*<br>CAN_DGN = 4242 (primary diagnosis code 4242), *or*<br>CAN_DGN = 4245 (primary diagnosis code 4245).<br><br>Reference:<br>4240 (diagnosis code) – PSC: Chron's Disease<br>4241 (diagnosis code) – PSC: Ulcerative Colitis<br>4242 (diagnosis code) – PSC: No Bowel Disease<br>4245 (diagnosis code) – PSC: Other Specify | Binary<br>1 indicates PSC | Static | – |
| Candidate Diagnosis of Primary Biliary Cholangitis<br>SAF Data Dictionary Code(s):<br>CAN_DGN = 4220 (primary diagnosis code 4220).<br><br>Reference:<br>4220 (diagnosis code) – Primary Biliary Cirrhosis (PBC) | Binary<br>1 indicates PBC | Static | – |
| Candidate Underwent Dialysis in the Prior Week<br>SAF Data Dictionary Code(s): CANHX_DIAL_PRIOR_WEEK | Binary<br>1 indicates "Yes" | Time-Varying | – |

**Table S.4:** The covariates associated with the *DynaMELD* model along with their respective encodings, temporal classifications, and units (where applicable). Each variable in this table is additionally annotated with the corresponding SAF Data Dictionary code(s) for reproducibility.

#### Appendix C.3 Missing Covariates and Imputation

| Column | Overall ($N$ = 53,046 $N_{obs}$ = 580,603) | *Training* ($N$ = 15,373 $N_{obs}$ = 212,873) | *Validation* ($N$ = 1,922 $N_{obs}$ = 27,760) | *Test* ($N$ = 1,922 $N_{obs}$ = 28,074) | *Pre-COVID Test* ($N$ = 7,513 $N_{obs}$ = 91,107) | *Post-COVID Test* ($N$ = 26,316 $N_{obs}$ = 220,789) |
|---|---|---|---|---|---|---|
| Sex | 0 (0.00%) | 0 (0.00%) | 0 (0.00%) | 0 (0.00%) | 0 (0.00%) | 0 (0.00%) |
| Serum Bilirubin | 244 (0.04%) | 73 (0.03%) | 5 (0.02%) | 9 (0.03%) | 45 (0.05%) | 112 (0.05%) |
| International Normalized Ratio | 245 (0.04%) | 73 (0.03%) | 5 (0.02%) | 9 (0.03%) | 46 (0.05%) | 112 (0.05%) |
| Serum Creatinine | 246 (0.04%) | 73 (0.03%) | 5 (0.02%) | 9 (0.03%) | 47 (0.05%) | 112 (0.05%) |
| Serum Sodium | 246 (0.04%) | 73 (0.03%) | 5 (0.02%) | 9 (0.03%) | 47 (0.05%) | 112 (0.05%) |
| Serum Albumin | 244 (0.04%) | 73 (0.03%) | 5 (0.02%) | 9 (0.03%) | 45 (0.05%) | 112 (0.05%) |
| Primary Diagnosis | 2782 (0.48%) | 101 (0.05%) | 0 (0.00%) | 0 (0.00%) | 135 (0.15%) | 2,546 (1.15%) |

**Table S.5:** The number (and percent) of missing entries associated with each covariate in our model covariate set, across the *training*, *validation*, *test*, *pre-COVID test*, and *post-COVID test* cohorts.

The covariates listed in Table S.5 are imputed using multiple imputation by chained equations (MICE). We use the implementation provided by scikit-learn[1]. Within this framework, we leverage a ridge regression model[2] to perform MICE imputation, and we restrict the imputed values to lie within the empirical minimum and maximum of its corresponding column. Features are imputed in order, starting with those containing the fewest missing values, and concluding with those containing the most missing values.

To impute missing data, we run MICE for either 10 iterations, or until the stopping criterion is reached; whichever is fewer. To formalize our stopping criterion, let $X_t$ represent the data matrix at the current iteration, $X_{t-1}$ represent the data matrix at the previous iteration, and $X_{obs}$ represent only the observed values in the data matrix. Then, our stopping criterion is whether the largest covariate change in the latest iteration, expressed in terms of the magnitude of values in the observed data, lies below a certain threshold. This is mathematically expressed as,

$$\frac{\max |X_t - X_{t-1}|}{\max |X_{obs}|} < 0.001.$$

To perform imputation, we fit a MICE model to the *training* set, then apply that fitted model to the other data splits without further fitting to those data. This is to avoid potential bias wherein the learned MICE model could impute the training data to match characteristics learned from the downstream evaluation sets (in our case, the *pre-COVID test* and *post-COVID test* sets). Since the data from these downstream evaluation sets are collected in the future, we do not want these characteristics to influence imputation of the training set.

As a final imputation step, we consider variables whose values are known to lie within a finite restricted set (e.g., the one-hot encoded diagnosis, which we know will lie within $\{0,1\}$), and we round imputed values to the nearest value within the set.

**Appendix C.4 Computing Time-to-Event/Censorship**

We calculate the time-of-event/censorship as follows. If the cause of a patient's removal from the waitlist was death, his or her time-of-event is:

$$t^{(i)} = t^{(i)}_{Death\ Date} - t^{(i)}_{Activation\ Date} - t^{(i)}_{Days\ from\ Listing}.$$

Taking the difference of the first two terms in the expression – the difference between the death and activation dates – computes the duration of the patient's waitlist tenure. Then, subtracting the number of days between the current patient entry and their date of listing computes the number of days between the current entry and the date of the patient's death.

If the patient was removed from the waitlist for any other reason, his or her time-of-event is similarly:

$$t^{(i)} = t^{(i)}_{Waitlist\ Removal\ Date} - t^{(i)}_{Activation\ Date} - t^{(i)}_{Days\ from\ Listing}.$$

In the above two equations, $t^{(i)}_{Days\ from\ Listing}$ is taken to be the difference (in days) between each candidate's date of waitlist record, and their waitlist activation date.

We calculate the event/censorship indicator for each patient as follows. Candidates who were removed from the waitlist due to medical unsuitability, death, or condition deterioration such that they became too sick to transplant are assigned an event indicator of 1, to indicate that they experienced the event (mortality or a suitable proxy). All other candidates are assigned an event indicator of 0, to indicate that they are right-censored.

Candidates with neither a date of removal nor date of death are assumed to correspond to those candidates occupying the liver transplant waitlist at the time of dataset collection. These instances are imputed with their last date of observation and are marked as right censored. Their time-of-censorship, $t^{(i)}$, is computed in the same way as for patients who were removed from waitlist for reasons other than pre-transplant mortality.

| SAF Data Dictionary Code | Encoding |
|---|---|
| Death Date<br>SAF Data Dictionary Code(s): CAN_DEATH_DT | Continuous |
| Activation Date<br>SAF Data Dictionary Code(s): CAN_ACTIVATE_DT | Continuous |
| Waitlist Removal Date<br>SAF Data Dictionary Code(s): CAN_REM_DT | Continuous |
| Date of Waitlist Record (Observation)<br>SAF Data Dictionary Code(s): CANHX_BEGIN_DT | Continuous |
| Cause of Removal<br>SAF Data Dictionary Code(s): CAN_REM_CD<br><br>Categories:<br>CAN_REM_CD = 5 (Candidate Medically Unsuitable)<br>CAN_REM_CD = 8 (Candidate Died on the Waitlist)<br>CAN_REM_CD = 13 (Candidate Condition Deteriorated) | Categorical |

**Table S.6:** Variables in the SRTR used to compute time-to-event/censorship and event indicators in our study. The variables in this table are annotated with SAF data dictionary codes for reproducibility.

##### Appendix C.5 Feature Normalization

Feature normalization refers to the statistical adjustments made to each covariate prior to passing it into the model. Empirical evidence suggests that normalizing the covariates in regression models improves model fit and generalization and avoids "exploding gradients" during learning.

In this study, we normalize our features in the following manner. Binary and categorical (one-hot encoded) covariates are not normalized. Left-skewed biological covariates – serum bilirubin, serum creatinine, and international normalized ratio – are *log*-normalized. This is accomplished by passing the natural logarithm of these values, rather than their raw quantities, into the model. Un-skewed biological covariates – serum sodium and serum albumin – and all other covariates are *z*-score normalized. If we use $x$ to denote the column in question, then *z*-score normalization applies the normalization equation,

$$x_{z-score} = \frac{x - \mu}{\sigma},$$

where $\mu$ represents the mean of the column $x$ in the training data, and $\sigma$ represents its standard deviation. Even in the *test*, *pre-COVID test*, and *post-COVID test* cohorts, we $z$-score normalize using the training mean and standard deviation, as in a deployment scenario, we would not have access to prospective summary statistics about future data that has yet to be gathered. Figure S.2 showcases the empirical distributions of each biological laboratory covariate before and after normalization.

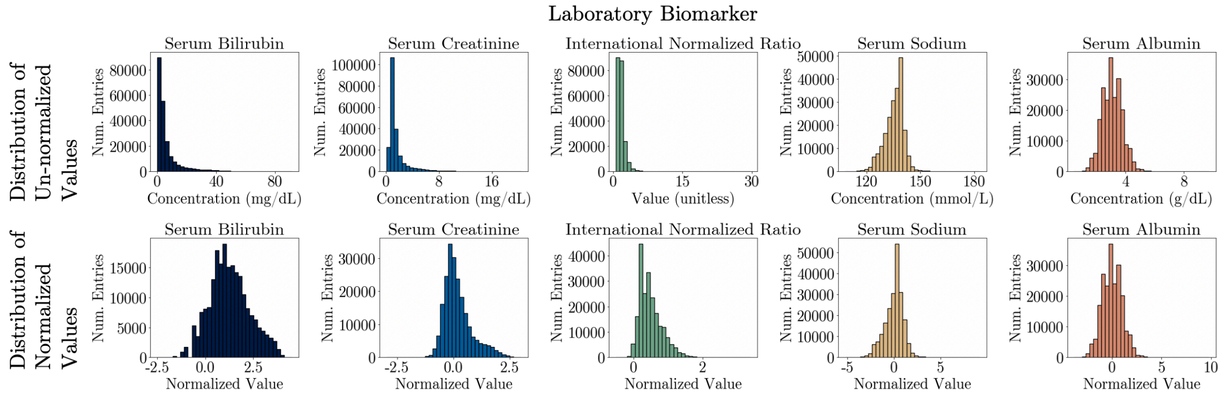

**Figure S.2:** Histogram plots showing the frequency of various un-normalized values of bilirubin, creatinine, INR, sodium, and albumin, respectively, in the *training* set (top row), and their normalized counterparts (bottom row). For each biomarker, these histograms plot the entry-wise frequency of each value (e.g., there may be more than one value per patient). Observe how *log*-normalization substantially reduces the left-skew of the un-normalized bilirubin, creatinine, and INR distributions. Furthermore, observe how the normalized values of these covariates lie close to zero: having covariates of small absolute value often prevents an explosion of gradients during model training.

In our study, rates-of-change of time-varying biological covariates are computed after normalization. The main body of this paper presents an example of computing the rate-of-change covariates associated with INR: hence, we discuss how to compute the rate-of-change of the *log*-INR, because INR is *log*-

normalized. However, the rate-of-change covariates corresponding to serum albumin and serum sodium are computed on the *z*-score normalized versions of those covariates.

### Appendix D: Model Implementation and Training

##### Appendix D.1: The DeepSurv Model

The DeepSurv model is an extension of Cox's proportional hazards model[3]. The Cox model aims to represents the hazard – the risk instantaneous risk of mortality/dropout if mortality/dropout has not yet occurred – associated with each instance in the data.

The Cox model decomposes the hazard into a "baseline hazard," which is a function only of time and can be estimated nonparametrically, and a "covariate-specific hazard" which depends only on instance covariates and is learned with gradient-based methods.

Under the Cox model, the hazard is represented as

$$h(t|x) = \lambda_0(t)\exp(\beta^T x),$$

where $\lambda_0$ is the baseline hazard function, $x \in \mathbb{R}^d$ represents $d$-dimensional patient covariates, $t \in \mathbb{R}_+$ represents time, and $\beta \in \mathbb{R}^d$ represents a set of model coefficients.

The DeepSurv architecture[4] relaxes this log-linear functional form used to represent the hazard under the Cox model. Under DeepSurv, the hazard is represented as

$$h(t|x) = \lambda_0(t)\exp\big(f_\theta(x)\big),$$

where $f_\theta \in \mathbb{R}^d \to \mathbb{R}$ is a neural network parameterized by $\theta$ mapping the $d$-dimensional covariate space to the real line. Because neural networks are universal function approximators[5,6], this formulation removes the Cox model's assumption that the hazard be a log-linear function of the instance covariates.

The parameters of both the Cox model and DeepSurv are trained by maximizing the Cox partial likelihood over the training data. For notation, assume we have a dataset of $D = \{(x^{(i)}, t^{(i)}, \delta^{(i)})\}_{i=1}^{N}$, where $x^{(i)}$ represents the covariates associated with patient $i$, $t^{(i)}$ represents the time of event (or right censorship), and $\delta^{(i)}$ is an indicator variable set to 1 if patient $i$ experiences the outcome of interest (death or removal from the waitlist) and 0 otherwise. Then, the optimization objective used to train DynaMELD can be written as

$$\theta^* = \arg\min_{\theta} \prod_{\{i:1,\ldots N\ s.t. \delta^{(i)}=1\}} \frac{\exp f_\theta(x^{(i)})}{\sum_{j \in R_{t^{(i)}}} \exp f_\theta(x^{(j)})} + \sum_{\theta' \in \theta} \|\theta'\|_2,$$

where $R_{t^{(i)}}$ represents the *risk set* at $t^{(i)}$, the set of patients who have neither died nor been censored as of $t^{(i)}$. The left-hand term of the above equation represents the standard Cox partial likelihood using a neural network ($f_\theta$), while the right-hand term represents a ridge regularization term over the network parameters.

**Appendix D.2: Optimization and Hyperparameters**

Each DynaMELD model was fit to the training data using the Adam optimizer[7] with validation-based early stopping[8] on 90-day concordance, with a 5-epoch patience threshold. The network weights are initialized using Kaiming initialization[9], which is the default initialization scheme for densely-connected neural network layers in PyTorch.

| Rate-of-Change Covariates | Number of Hidden Layers | Size of Hidden Layers | $\ell_2$ Regularization Coefficient | Learning Rate |
|---|---|---|---|---|
| None<br>First-Order ($\Delta$)<br>First- and Second-Order ($\Delta, \Delta^2$) | 0 (Cox Model)<br>1<br>3<br>5 | 128<br>256<br>512<br>1024 | $1 \times 10^{-3}$<br>$1 \times 10^{-2}$ | $1 \times 10^{-3}$<br>$1 \times 10^{-2}$ |

**Table S.7:** Table of hyperparameters explored during optimization. We trained one DynaMELD model for each combination of entries in the above columns. In our results, for each set of features and rate-of-change covariates, we report only the performance associated with the model that achieves the highest 90-day concordance on the *validation* set.

##### Appendix D.3: Model Output and Rescaling

To allow practitioners to draw upon their clinical intuition when working with DynaMELD, we rescale the score so that the risk percentile associated with a given DynaMELD score matches that of the MELD 3.0. Put differently, a clinicians' intuition regarding relative risk should be invariant between DynaMELD and the MELD 3.0: a patient assigned a DynaMELD score of 30, for example, will be at greater risk than the same proportion of the population as a patient assigned a MELD 3.0of 30.

We implement this percentile matching as follows. We compute the empirical cumulative density of risk scores assigned to patient instances under both the MELD 3.0 ($F_{MELD3.0}$) and DynaMELD[‡] ($F_{DynaMELD}$) on each entry of the *training* set, at a granularity of $5 \times 10^{-5}$. Then, for each instance's DynaMELD score, $y^{(i)}_{DynaMELD}$, we find its corresponding MELD 3.0 by first calculating the quantile of $y^{(i)}_{DynaMELD}$ under DynaMELD, then finding the MELD 3.0 score corresponding to that same quantile under the MELDNa. Mathematically, this procedure is expressed using the following two equations:

$$Quantile\left(y^{(i)}_{DynaMELD}\right) = F_{DynaMELD}\left(y^{(i)}_{DynaMELD}\right),$$

$$y^{(i)}_{DynaMELD-rescaled} = F^{-1}_{MELD3.0}\left(Quantile\left(y^{(i)}_{DynaMELD}\right)\right).$$

#### Appendix E: Baselines and Evaluation

‡ Here, "DynaMELD" refers to the individual variant of DynaMELD that is being rescaled. We perform this same procedure once for each DynaMELD model.

**Appendix E.1: Computing Comparison Baselines**

In this section, we provide the equations used to compute the comparison baselines of MELD, MELDNa, and MELD 3.0. The notation $a|_{[b,c]}$ denotes restricting $a$ to lie within the range $[b,c]$. The notation $a|_{[b,\ldots,c]}$ denotes rounding $a$ to the nearest integer within the range $[b,c]$. In each case, square brackets denote an inclusive range boundary, while round brackets denote an exclusive range boundary. Following how the MELDNa and MELD 3.0 are computed by UNOS for prioritization, we set a patient's level of serum creatinine to 4 mg/dL (for MELDNa), and to 3 mg/dL (for MELD 3.0) if the patient has undergone dialysis treatment in the past week.

$$\begin{aligned}\mathrm{MELD}(X) = \Big[&9.57\log\left(X_{\mathrm{creatinine}}^{(\mathrm{mg/dL})}|_{[1,4]}\right)\\ &+ 3.78\log\left(X_{\mathrm{bilirubin}}^{(\mathrm{mg/dL})}|_{[1,\infty)}\right) + 11.2\log\left(X_{\mathrm{INR}}|_{[1,\infty)}\right) + 6.43\Big]\,|_{[6,\ldots,40]}\end{aligned}$$

$$\mathrm{MELDNa}(X) = \left[\mathrm{MELD}(X) + 1.59\left(135 - X_{\mathrm{sodium}}^{(\mathrm{mmol/L})}|_{[125,137]}\right)\right]|_{[6,\ldots,40]}$$

$$\begin{aligned}\mathrm{MELD3.0}(X) = \Big[&1.33X_{\mathrm{female}} + 4.56\log\left(X_{\mathrm{bilirubin}}^{(\mathrm{mg/dL})}|_{[1,\infty)}\right)\\ &+ 0.82\left(137 - X_{\mathrm{sodium}}^{(\mathrm{mmol/L})}|_{[125,137]}\right)\\ &- 0.24\left(137 - X_{\mathrm{sodium}}^{(\mathrm{mmol/L})}|_{[125,137]}\right)\log\left(X_{\mathrm{bilirubin}}^{(\mathrm{mg/dL})}|_{[1,\infty)}\right)\\ &+ 9.09\log\left(X_{\mathrm{INR}}|_{[1,\infty)}\right) + 11.14\log\left(X_{\mathrm{creatinine}}^{(\mathrm{mg/dL})}|_{[1,\infty)}\right)\\ &+ 1.85\left(3.5 - X_{\mathrm{albumin}}^{(\mathrm{g/dL})}|_{[1.5,3.5]}\right)\\ &- 1.83\left(3.5 - X_{\mathrm{albumin}}^{(\mathrm{g/dL})}|_{[1.5,3.5]}\right)\log(X_{\mathrm{creatinine}}^{(\mathrm{mg/dL})}|_{[1,\infty)})\\ &+ 6\Big]\,|_{[6,\ldots,\infty)}\end{aligned}$$

#### Appendix E.2: 90-Day Concordance Index

We compute 90-day concordance by censoring all patients with $t^{(i)} > 90$ (assigning such patients a surrogate event indicator value of 0), then computing Harrell's concordance statistic over this modified dataset. We use the implementation of Harrell's concordance index provided by the `lifelines` Python package[10].

For the sake of subsequent comparison against the Pooled Group Concordance Index (Appendix E.3), we now discuss the computation of Harrell's concordance index and the quantity that it represents. One common means of constructing Harrell's concordance estimator is as a ratio of the cardinality of two sets. The *comparable set* is the set of all pairs of patients for whom we can determine, from their times of event/censorship, which member of the pair first experienced pre-transplant mortality. This is equivalent to the set of pairs for which the member of the pair with the lowest time-of-event/censorship is uncensored. The *concordant set* represents the subset of comparable pairs for which a risk model, $f$, correctly identifies the patient who experiences the event first. Harrell's concordance index is the ratio between the number of pairs in the concordant set and the number of pairs in the comparable set.

$$S_{Comparable} = \{(i,j) \mid t^{(i)} < t^{(j)}, \delta^{(i)} = 1\}$$

$$S_{Concordant} = \left\{(i,j) \in S_{Comparable} \,\middle|\, f\left(x^{(i)}\right) > f\left(x^{(j)}\right)\right\}$$

$$C_{Harrell} = \frac{|S_{Concordant}|}{|S_{Comparable}|}$$

Semantically, Harrell's concordance index represents the fraction of patient pairs who are well-ordered under a risk model (e.g., the patient with the higher risk score empirically dies first in the observed data). The index ranges from 0 and 1: a score of 1 indicates perfect relative risk discrimination, a score of 0.5 indicates the expected concordance under random risk assignment, and a score of 0 indicates perfect inverse concordance.

#### Appendix E.3: Pooled Group 90-Day Concordance Index and Identifying Subpopulations

The Pooled Group Concordance Index (PGCI) provides one means of evaluating the fairness of a survival model[11]. In addition to taking as input each patient's time-of-event/censorship and event indicator, the PGCI also accepts an indicator variable, $a_i$, identifying whether each patient is a member of a certain identified sub-group of interest (e.g., $a_i = 1$ if patient $i$ is female; otherwise, $a_i = 0$). The PGCI modifies Harrell's concordance index by requiring that the first member of each pair in $S_{Comparable}$ also be a member of the identified sub-group. Computation of the PGCI is governed by the following equations.

$$S_{Comparable,a} = \{(i,j) \mid t^{(i)} < t^{(j)}, \delta^{(i)} = 1, a^{(i)} = 1\}$$

$$S_{Concordant,a} = \left\{(i,j) \in S_{Comparable,a} \mid f\left(x^{(i)}\right) > f\left(x^{(j)}\right)\right\}$$

$$C_{Pooled\ Group,a} = \frac{|S_{Concordant,a}|}{|S_{Comparable,a}|}$$

Semantically, the PGCI quantifies how well a given risk score predicts the relative risk of members of the identified sub-group compared to that of the rest of the patient population.

Although it is an intuitive statistic to measure the fairness of survival regression models, the primary limitation of PGCI is that it represents a *demand-side* measure of inequity – it does not account for *supply-side* factors that may disadvantage certain sub-groups.

Demand-side inequity refers to sources of inequity resulting from the level of risk assigned to each patient on the waitlist. As an example, consider how the MELDNa score contains a positive coefficient of serum creatinine, yet does not adjust for patient sex. Because serum creatinine is positively correlated with muscle mass, and because male patients tend to have a higher degree of muscle mass than female patients, this suggests that a male patient with a fixed degree of liver dysfunction would be assigned a higher MELDNa than his similarly sick female counterpart. Such a source of inequity would be well-captured by the PGCI, because a risk score with this bias would not accurately rank the risk of female patients relative to the total population. In fact, we hypothesize that the MELD 3.0's adjustment for patient sex explains much of its improvement over the MELDNa in PGCI when the identified sub-group corresponds to female patients.

Supply-side inequity refers to sources of inequity that stem from unequal allocation of organs to similarly sick patients awaiting transplant. As an example, consider how the body habitus of different patients influences their ability to attract deceased-donor organs for transplant. All else being equal, a patient with a smaller body habitus enjoys a smaller pool of compatible livers than a patient with a larger physical build. In general, this makes it easier for male patients, who tend to be physically larger, to attract deceased-donor livers for transplant than their female counterparts, who tend to be smaller. However, as the magnitude of this bias is not determined by how accurately female patients are ranked relative to their male counterparts, this form of bias would not be well-captured under the PGCI.

In this work, we primarily focus on studying fairness with respect to women, patients with primary sclerosing cholangitis, and patients with primary biliary cholangitis. The specific selection criteria that we used to identify members of these sub-groups for study can be found below in Table S.11. These same selection criteria are applied to other cases where we wish to identify members of these sub-groups, such as when computing quantile reclassification statistics (Appendix E.4).

| Identified Sub-Group | Identification Criterion |
|---|---|
| Women | Patient Sex is Female<br>Criterion:<br>CAN_GENDER = 1 (Female) |
| Patients with Primary Sclerosing Cholangitis | Patient's Diagnosis Code Corresponds to Primary Sclerosing Cholangitis<br>Criterion:<br>CAN_DGN = 4240 (Primary Sclerosing Cholangitis, Chron's Disease), *or*<br>CAN_DGN = 4241 (Primary Sclerosing Cholangitis, Ulcerative Colitis), *or*<br>CAN_DGN = 4242 (Primary Sclerosing Cholangitis, No Bowel Disease), *or*<br>CAN_DGN = 4245 (Primary Sclerosing Cholangitis, Other Specify) |

| | |
|---|---|
| Patients with Primary Biliary Cholangitis | Patient's Diagnosis Code Corresponds to Primary Biliary Cirrhosis<br>Criterion:<br>CAN_DGN = 4220 (Primary Biliary Cirrhosis) |

**Table S.8:** The specific selection criteria, annotated with SAF (Standard Analysis File) Data Dictionary codes for reproducibility, that we used to identify sub-groups for this study.

**Appendix E.4: Quantile Reclassification**

Quantile reclassification provides an intuitive means to assess the comparative performance of survival models on sub-populations of interest. The underlying argument is as follows: if we can identify a sub-population who was under-served by the status quo, and we can demonstrate that the proposed model places these patients into a higher risk percentile (quantile) than the current standard – which would afford these patients a higher chance of attracting a deceased-donor transplant liver during their listing period – then the proposed model represents a superior approach for the given sub-population.

Our specific procedure for this analysis is as follows. We first extract each patient's first observation from evaluation dataset (e.g., *pre-COVID test* set), and calculate the corresponding risk assigned by each model (MELDNa, MELD 3.0, DynaMELD) on each entry. Then, for each risk score (e.g., DynaMELD), we compute the empirical cumulative density of its assigned risks ($F_{model}$), and use this to compute the risk quantile associated with of each entry. Mathematically, for a given patient $i$, this is expressed as follows:

$$Quantile\left(y_{model}^{(i)}\right) = F_{model}\left(y_{model}^{(i)}\right)$$

Each patient $i$ is then placed into one of five bins, depending on the value of $Quantile\left(y_{model}^{(i)}\right)$. This is shown below in Table S.12. By performing this binning, we obtain the below $5 \times 5$ matrix where the rows highlight each patient's risk percentile under MELD 3.0, and the columns highlight each patient's risk percentile under DynaMELD. Observe that the upper-right corner of this matrix represents patients who are moved into a higher risk tier under DynaMELD, while the lower-left corner represents those who are moved into a lower tier. The diagonal represents patients who risk percentile did not significantly change between the two scores. The number of *up-ranked* patients is therefore calculated by summing the upper-right corner of this matrix, while the number of *down-ranked* patients is calculated by summing the lower-left corner. By performing this procedure 1,000 times and resampling (with replacement) the dataset each time, we obtain bootstrapped confidence intervals on the number of up- and down-ranked patients assigned under each score.

| Reclassification Quantiles, All **Decedents** in *Pre-COVID Test* Cohort ($n = 1{,}340$) | | | | | | |
|---|---|---|---|---|---|---|
| | | Risk Percentile Under DynaMELD, n (%) | | | | |
| | | 0 – 20 (Lowest Risk) | 20 – 40 | 40 – 60 | 60 – 80 | 80 – 100 (Highest Risk) |
| Risk Percentile Under MELD 3.0, n (%) | 0 – 20 (Lowest Risk) | 140 (10.5%) | 70 (5.2%) | 22 (1.7%) | 0 (0.1%) | 0 (0.0%) |
| | 20 – 40 | 36 (2.7%) | 113 (8.5%) | 62 (4.7%) | 9 (0.7%) | 0 (0.0%) |
| | 40 – 60 | 3 (0.2%) | 68 (5.1%) | 159 (11.9%) | 97 (7.3%) | 2 (0.2%) |
| | 60 – 80 | 0 (0.0%) | 1 (0.1%) | 34 (2.6%) | 181 (13.6%) | 43 (3.2%) |
| | 80 – 100 (Highest Risk) | 0 (0.0%) | 0 (0.0%) | 0 (0.0%) | 31 (2.3%) | 260 (19.4%) |
| Up-ranked: 309.0 (95% CI: 277.0, 339)<br>Down-ranked: 175.0 (95% CI: 152.0, 200)<br>Difference (num. more up-ranked): 134.0 (95% CI: 92.0, 177.0); 10.0% of decedents. | | | | | | |

**Table S.9:** Table highlighting the quantile reclassification procedure that we employ to study the number of patients placed into higher and lower risk tiers under DynaMELD compared to baseline risk scores. This example highlights how decedents in the *pre-COVID test* cohort patients would be reclassified under DynaMELD compared against the MELD 3.0. While the individual cells in the table contain results from only a single resampled version of the *pre-COVID test* cohort, the confidence intervals associated with the number of up- and down-ranked patients are computed from 1,000 bootstrap resamples.

Typically, sub-populations studied in this context are identified by being groups of patients who have uniquely high historical rates of pre-transplant mortality relative to the general waitlisted population. One such population is the set of patients who died prior to receiving an organ under the status quo (MELDNa): clearly, this group has the highest rate of historical pre-transplant mortality relative to the general waitlisted population. Because these patients did not attract a deceased-donor organ in time, they were evidently under-served by the existing risk score and would have benefitted from being placed in a higher risk quantile during their listing. However, decedents are not the only sub-population worthy of study under this method: we can draw on existing literature that identifies sub-populations with higher rates of pre-transplant mortality, such as women, and perform the same analysis.

One advantage of this method is that, depending on how sub-populations for study are identified, reclassification statistics can highlight the supply-side inequities discussed in Appendix E.2 that may not be apparent under the PGCI. Because the historical rate of pre-transplant mortality in a population depends upon both supply and demand-side factors, using historical pre-transplant mortality as a selection criterion allows us to determine the extent to which our approach corrects for inequities that result from both supply-side and demand-side factors. However, the principal limitation of this method is that it cannot tell us the desired magnitude of reclassification, only the direction. Put differently, it may be possible to over-correct with respect to a specific sub-group (e.g., to raise the risk quantiles of a particular sub-groups to the point where it becomes unfair to other sub-groups), and this method is incapable of distinguishing an over-corrected adjustment from a well-calibrated one.

Our analysis comes with two additional caveats. First, although we compare quantile reclassification statistics of DynaMELD against the MELD 3.0, the empirical data identifying the relative disadvantage of each sub-group was generated under MELDNa-based allocation. It is not known whether each decedent under MELDNa-based allocation would remain a decedent under the MELD 3.0, as it is possible that some of the identified decedents under MELDNa may not have died under MELD 3.0-based allocation. Although our results include results comparing the reclassification of patient sub-groups under DynaMELD vs. MELD 3.0, these results are merely intended to give a picture of how different sub-populations may fare under each allocation system. In such results comparing DynaMELD vs. MELD 3.0, a larger number of up-ranked patients should <u>not</u> be taken to imply a "fairer" algorithm.

Second, to avoid biasing the reclassification statistics in favour of patients with more observed instances, each patient's risk (and subsequent reclassification) is computed based on his or her record collected at listing. However, in the context of a dynamic risk score like DynaMELD, this may be limiting, as such data does not let us readily assess the influence of rate-of-change covariates on performance (see F.2. for further discussion of this limitation). We leave to future work the determination of a reclassification statistic that appropriately reflects the dynamic nature of each patient's waitlist tenure.

### Appendix F: Additional Experiments and Results

##### Appendix F.1: Concordance Results Across Data Partitions

Figure 3 in the main body presents only the 90-day concordance and 90-day pooled group concordance on the *pre-COVID test* split of the data. In this section, we additionally provide plots highlighting those same concordance results on the *test* and *post-COVID test* partitions.

*Appendix F.1.1: Main Concordance Results, Test Split*

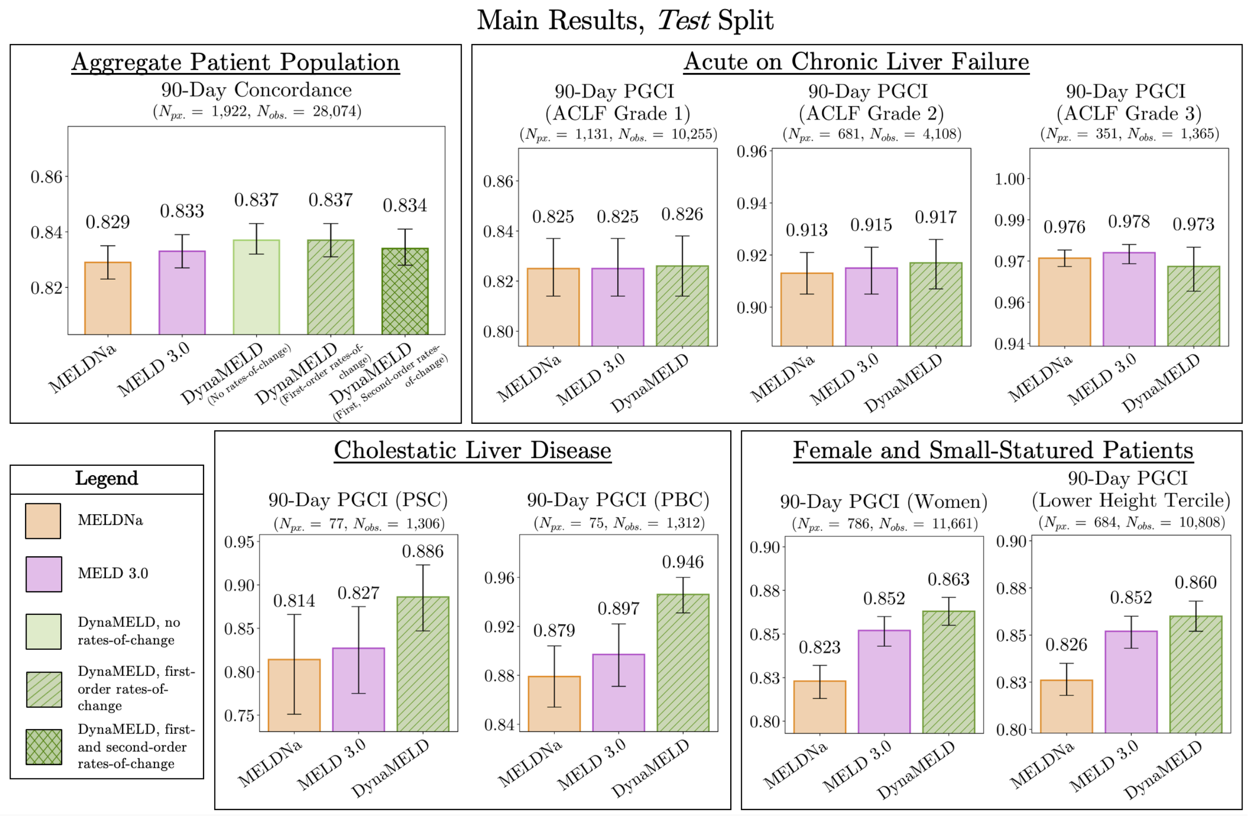

**Figure S.3:** 90-day concordance and PGCI statistics computed across each entry of the *test* set. Observe how the performance of the baselines and of DynaMELD on these two datasets is like that on the *pre-COVID test* set presented in the main manuscript.

*Appendix F.1.2: Main Concordance Results, Post-COVID Test Split*

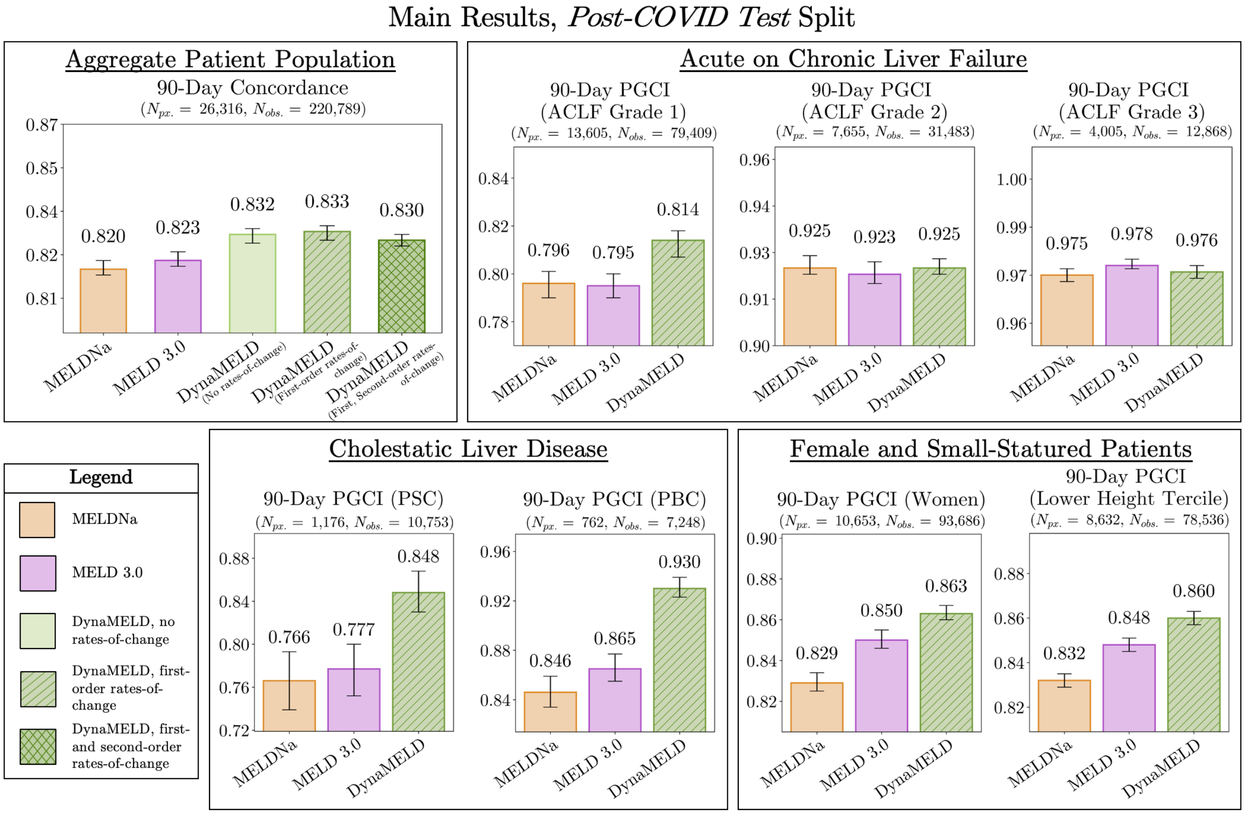

**Figure S.4:** 90-day concordance and PGCI statistics computed across each entry of the *post-COVID test* set. Observe how the performance of the baselines and of DynaMELD on these two datasets is like that on the *pre-COVID test* set presented in the main manuscript.

**Appendix F.2: Relative Risk Discrimination**

The analysis presented above and in the main body of this paper assesses 90-day concordance (and pooled group 90-day concordance) *on each entry* in the data. This was a deliberate decision, as we wanted to assess the performance of each model at a time that is largely agnostic to when the patient was listed. This means that if patient $i$ dies pre-transplant, and has multiple observations, our 90-day concordance statistic will treat pairs of observations with both entries corresponding to patient $i$ as separate, comparable pairs.

One advantage of this approach is that it allows us to better capture the implicit assumption that each patient's risk should grow over time, as each patient approaches his or her time-to-pre-transplant-mortality. However, as there are considerably more entries than patients (Appendix B.2), this approach may be biased to favour methods that perform better patients with more observations. In this section, we discuss two alternatives to entry-wise 90-day concordance that would not suffer from this form of bias; each provides an alternative means of obtaining one entry per patient prior to computing concordance.

1. <u>90-Day Concordance (From Listing)</u>. This approach entails computing concordance over the patient population based on each patient's assigned risk at listing. To our knowledge, this is how concordance is computed in the evaluation of the MELD, MELDNa, and MELD 3.0. Because this approach leverages only one observation per patient, it does not bias the metric in favour of patients with more observations. There are, however, two limitations to this approach. First, because each patient's record at listing is their first record, these observations do not contain the historical biomarker trajectories that are leveraged in our dynamic score. Therefore, this approach does not let us readily assess the influence of rate-of-change covariates on predictive performance: any variation in 90-day concordance from listing due to including rate-of-change covariates would be mediated by the indirect effects of those covariates on the training dynamics of the model. Second, this statistic is highly dependent on the time at which each patient is added

to the waitlist, and so may not generalize well to regions or countries in which patients are listed substantially earlier or later than those in the OPTN cohort.

2. 90-Day Concordance (Random Sample). This approach entails sampling one record for each patient in the cohort and computing 90-day concordance using these subsampled records. While this approach does not bias our metric in favour of patients with more observations, it does not capture the assumption that each patient's risk of mortality should increase as a function of time. Furthermore, this approach may be sensitive to the label error of the OPTN dataset discussed in the main manuscript.

*Appendix F.2.1: Main Concordance Results, At Listing*

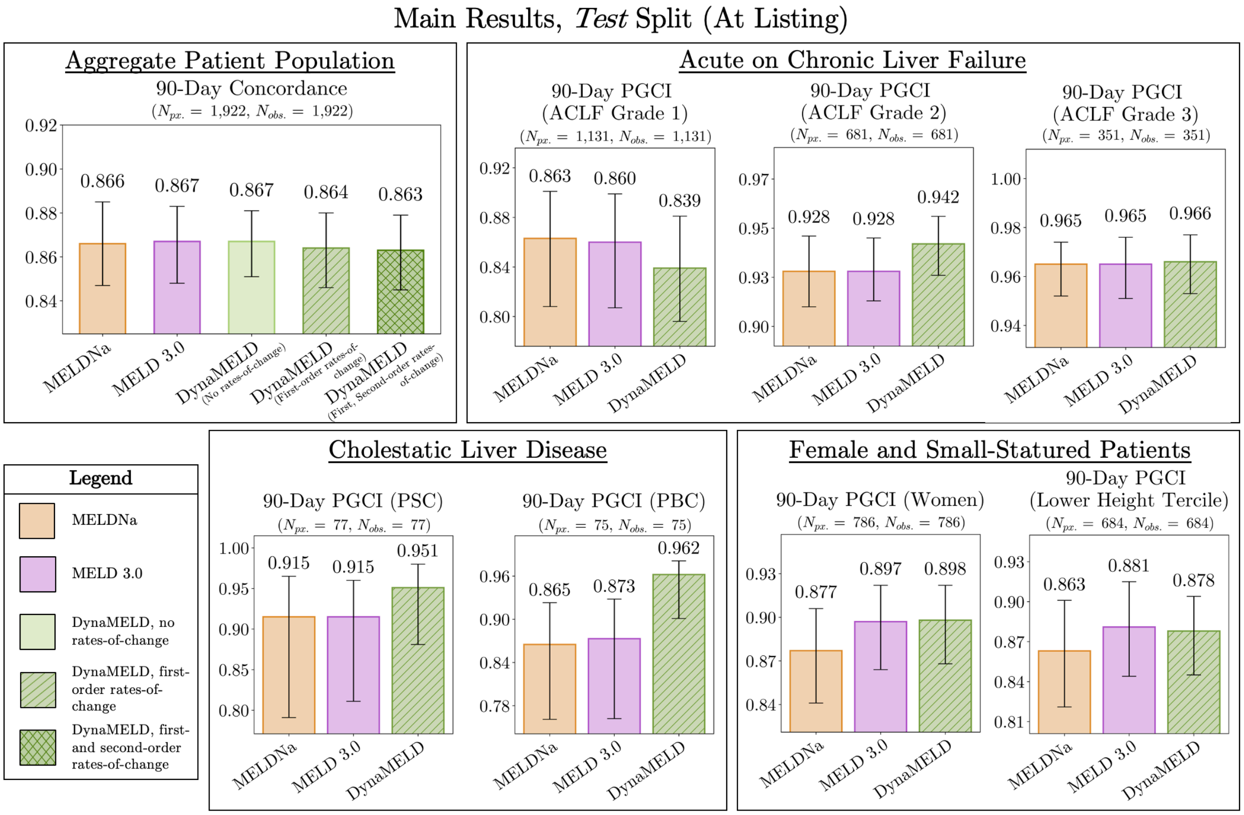

Main Results, *Pre-COVID Test* Split (At Listing)

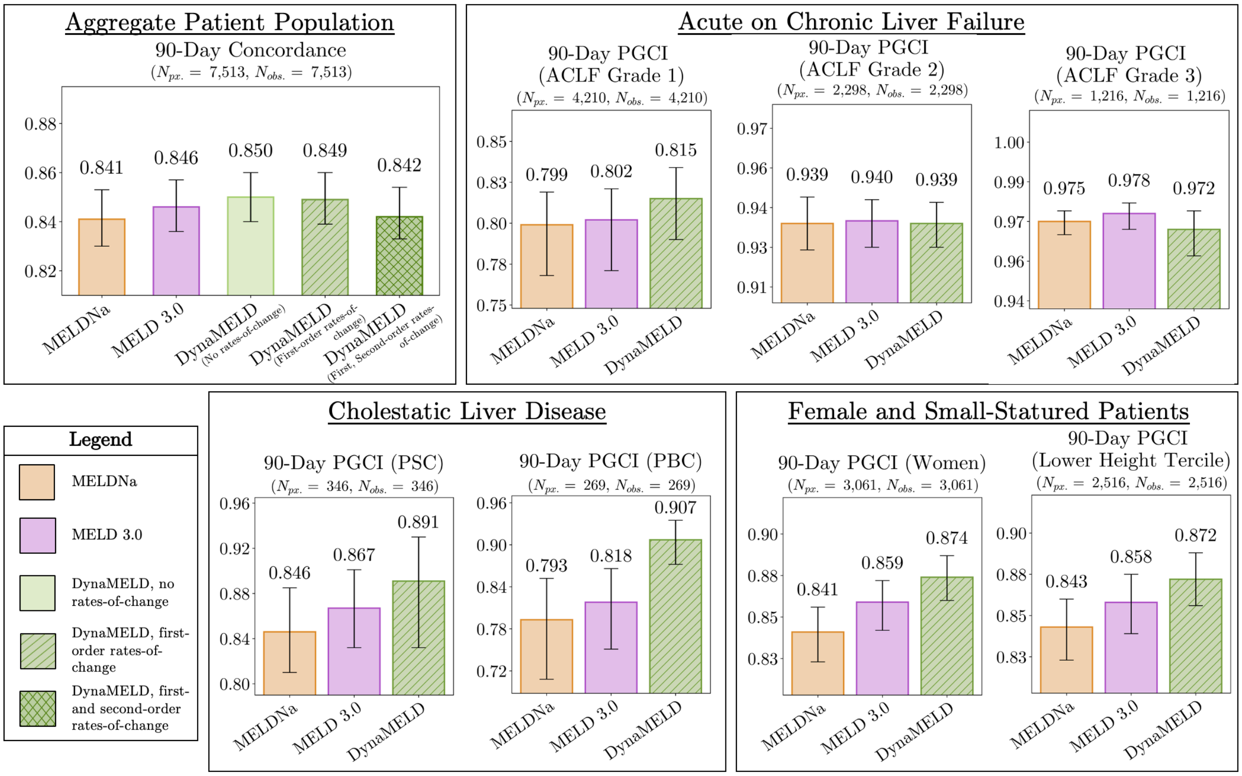

Main Results, *Post-COVID Test* Split (At Listing)

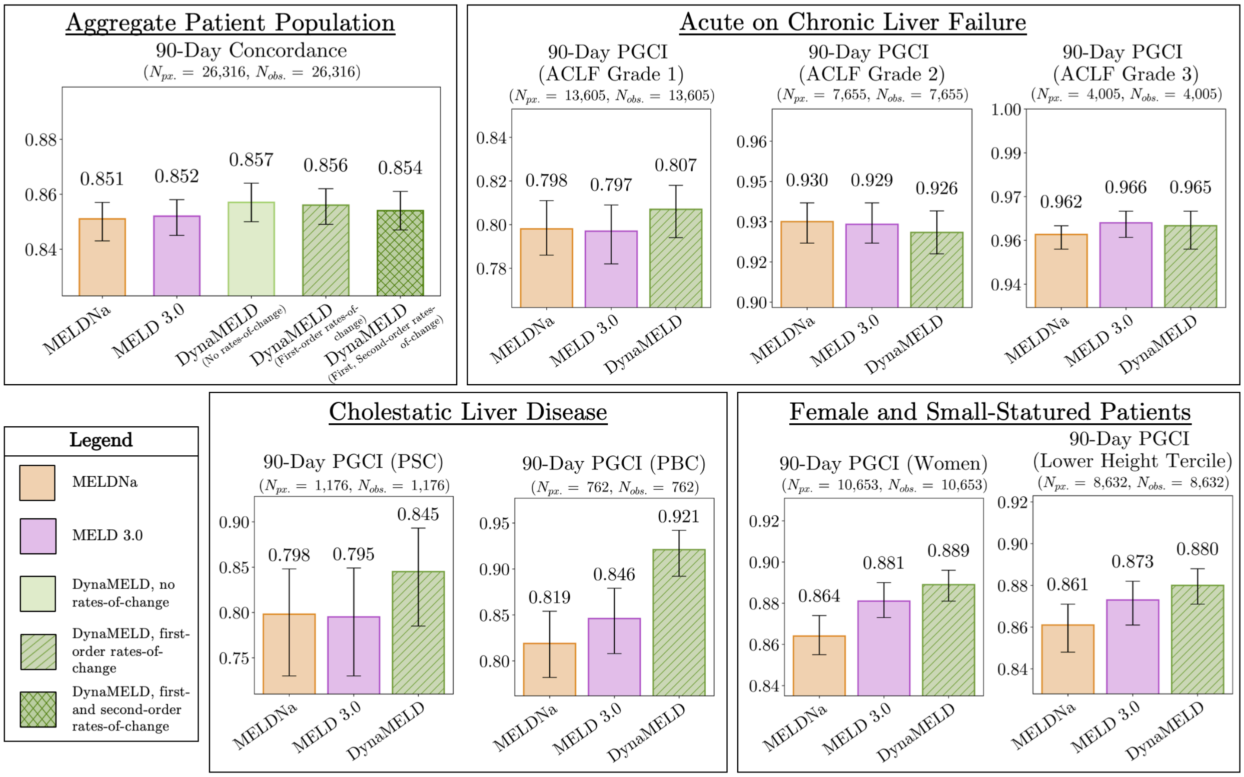

**Figure S.5:** 90-day concordance statistics, computed from the time of patient listing on the *test*, *pre-COVID test*, and *post-COVID test* sets. Observe how DynaMELD continues to outperform the baselines on PGCI in this setting, and achieves comparable performance on 90-day concordance. Additionally, observe how the concordance statistics of the MELDNa and MELD 3.0 on the *test* set, 0.866 and 0.867, respectively, more closely match the concordance statistics reported in the MELD 3.0 paper (0.869 and 0.862, respectively)[12]. From there, however, some decline in 90-day concordance at listing is observed across all models on the *pre-COVID test* and *post-COVID test* sets.

*Appendix F.2.1: Main Concordance Results, Random Sample*

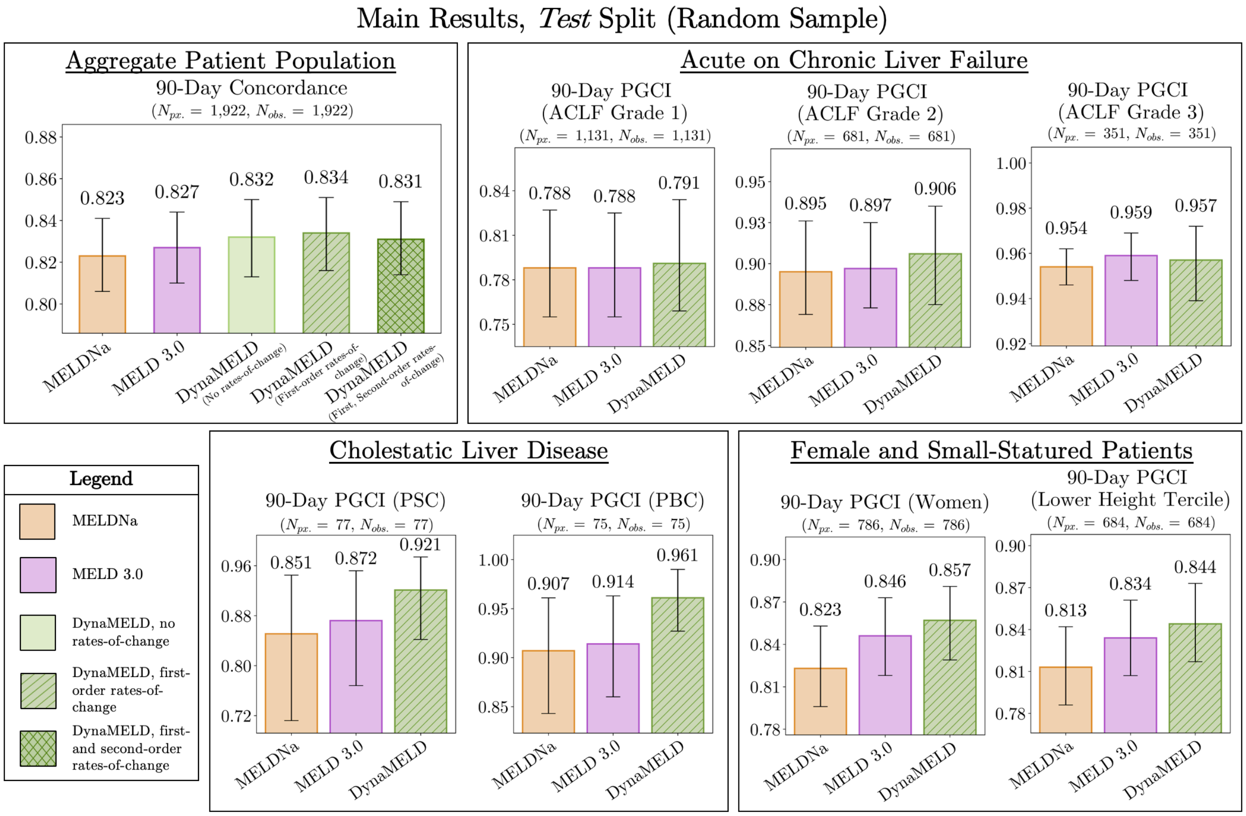

Main Results, *Pre-COVID Test* Split (Random Sample)

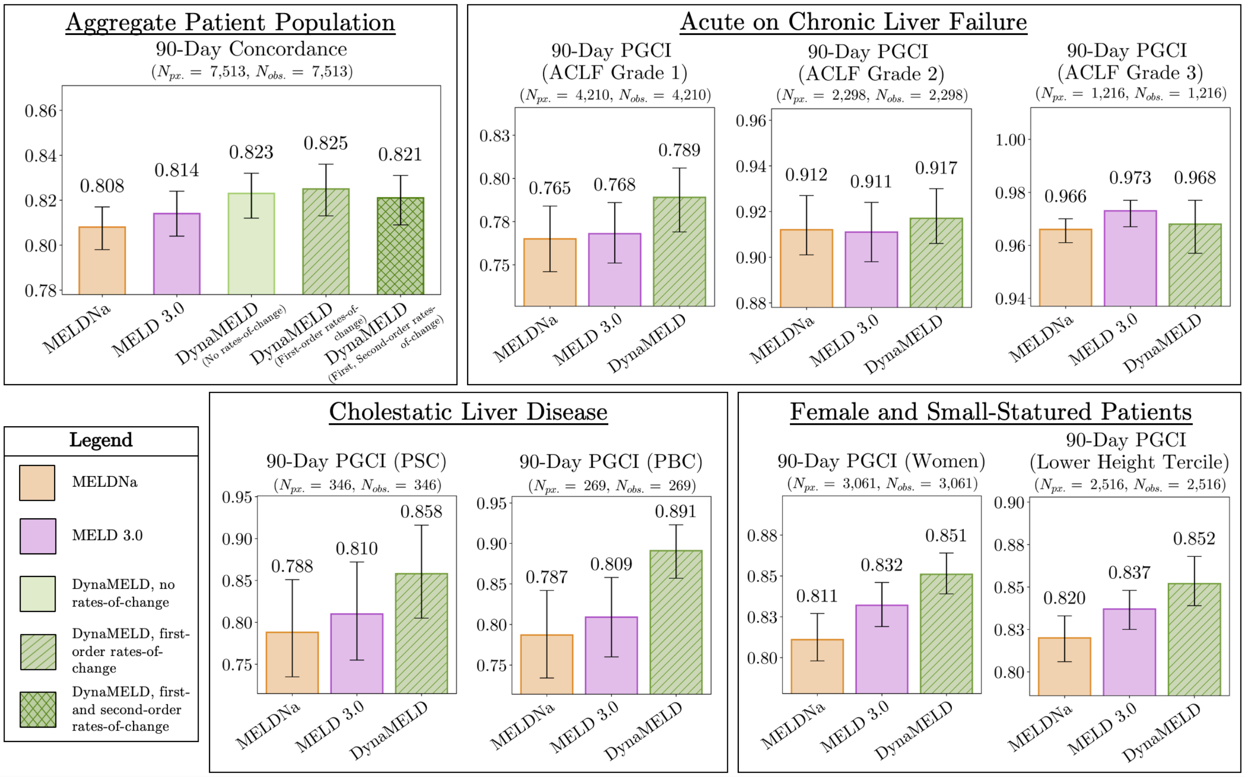

Main Results, *Post-COVID Test* Split (Random Sample)

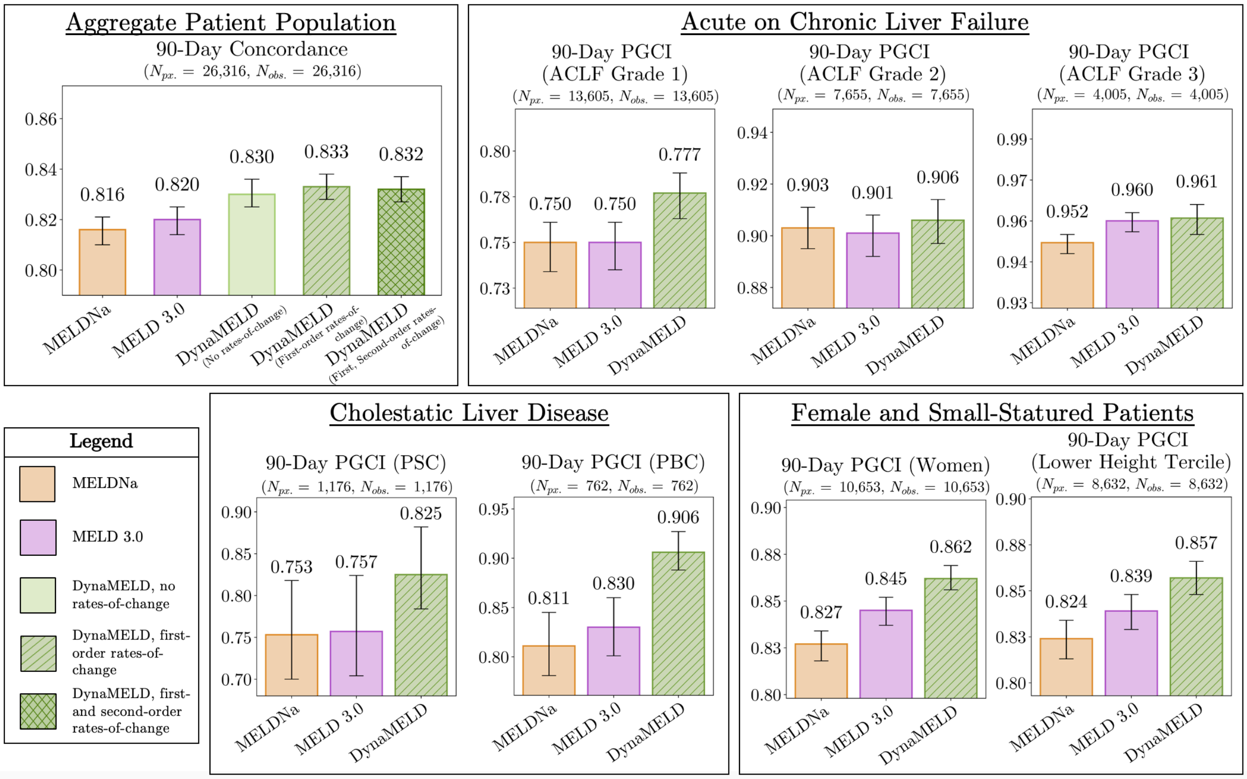

**Figure S.6:** 90-day concordance statistics, computed across a random observation sampled from each patient listed in the *test*, *pre-COVID test*, and *post-COVID test* sets. Observe how DynaMELD (with few exceptions) outperforms the baselines in this setting.

**Appendix F.3: Robustness to Regional Variation**

Organ allocation in the United States is practiced at the level of OPTN *allocation regions* – with few exceptions, donor organs from a given region are matched to recipients within the same region.

DynaMELD, like the MELD, MELDNa, and MELD 3.0, is trained on patient data aggregated across regions. This training scheme admits a possible aggregation bias, wherein the model learns to correctly rank patients in the aggregate population by relative risk but achieves poor generalization within each region. In this section, we present concordance results associated with each OPTN allocation region to study the extent to which such bias is present under our model. Here, we report results on the *pre-COVID test* cohort.

Table S.10 provides summary statistics regarding the number of patients and entries in each region in the *pre-COVID test* set. Figures S.7 and S.18highlight the 90-day concordance achieved by DynaMELD and each of the baselines within each of the 11 OPTN regions. As in our main results, concordance was computed over each patient observation in the data. Overall, this plot shows that, while some extent of aggregation bias is present, they do not substantially impact the main results. Although there are regions within which DynaMELD does not outperform the baselines, the general trend across regions shows that DynaMELD improves relative risk discrimination. The presence of aggregation bias in these findings highlights the need for future work to explore the performance of region-specific models, compared to that of models trained across the entire nationwide patient cohort.

| OPTN Region Number | Number of Patients (*Pre-COVID Test Set*) | Number of Entries (*Pre-COVID Test Set*) |
|---|---|---|
| 1 | 437 | 7,290 |
| 2 | 905 | 10,778 |
| 3 | 1,176 | 11,107 |
| 4 | 835 | 10,326 |
| 5 | 1,160 | 16,014 |
| 6 | 203 | 2,611 |
| 7 | 563 | 7,591 |
| 8 | 388 | 3,761 |
| 9 | 441 | 7,173 |
| 10 | 701 | 6,336 |
| 11 | 702 | 8,107 |
| *Total* | 7,513 | 91,107 |

**Table S.10:** Number of patients and entries associated with each OPTN region in the *pre-COVID test* set. In Figures S.9, observe how regions with fewer patients and instances tend to yield wider confidence intervals.

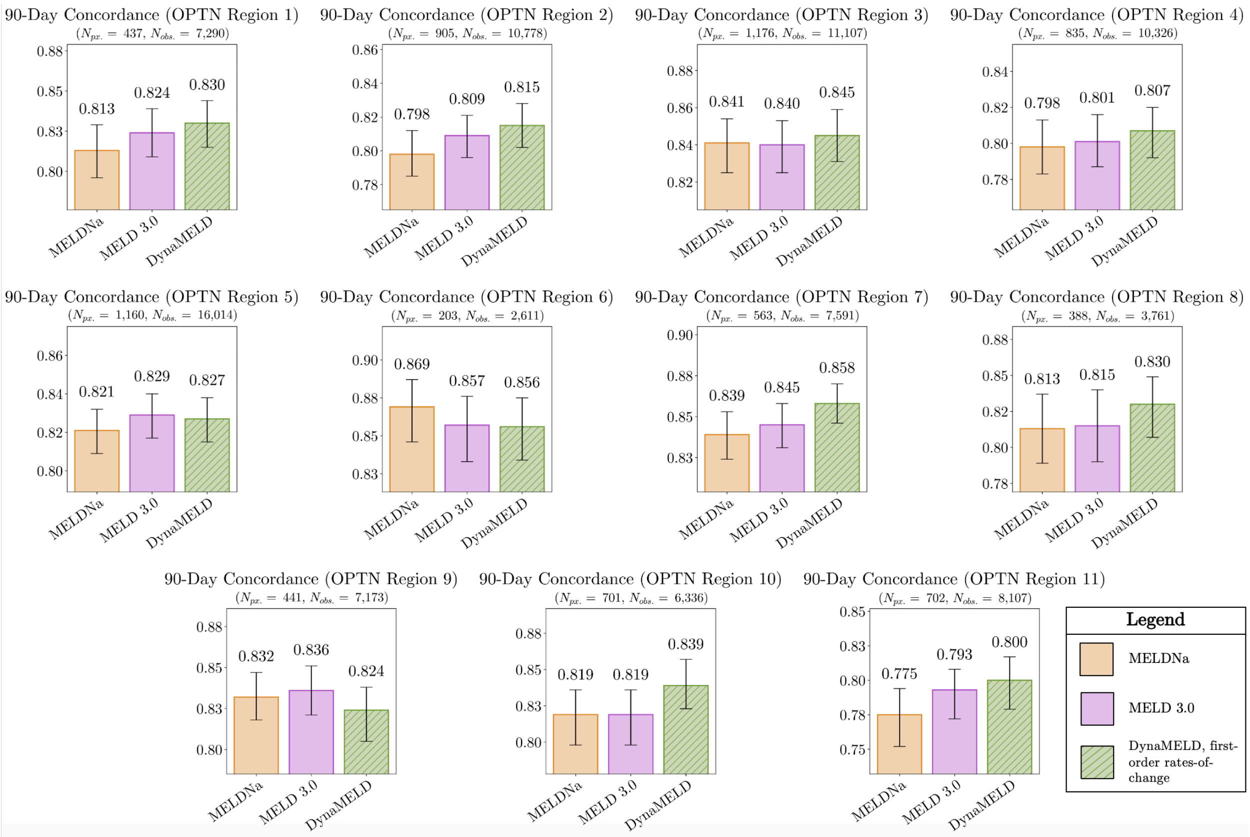

**Figure S.7:** 90-day pooled group concordance statistics computed separately over each of the 11 OPTN regions. These concordance statistics were calculated over all instances in each observed region. Although these results suggest some degree of aggregation bias is incurred by our training scheme, DynaMELD obtains higher 90-day concordance than the baseline models in all settings except for Regions 5, 6, and 9.

**Appendix F.5: Characterizing Acute-on-Chronic Liver Failure**

To abide by the EASL criteria for characterizing acute-on-chronic liver failure as closely as is possible within the constraints of the SRTR data repository, we adopt the following scheme.

A patient is assigned one renal point if he or she underwent dialysis in the previous week (CANHX_DIAL_PRIOR_WEEK) at the time in question, or if their measured level of serum creatinine (CANHX_SERUM_CREAT) at the time in question is greater than 2 mg/dL. Otherwise, zero renal points are assigned. A patient is assigned one respiratory point if they are on a ventilator at *any* time point during their listing (CAN_VENTILATOR); while it would be preferable to assign these points only at the time point in question, the CAN_VENTILATOR variable is recorded in the SRTR data repository only once for each patient (and <u>not</u> for each time point). Otherwise, zero respiratory points are assigned. Similarly, a patient is assigned one circulatory point if the patient is on life support (CAN_LIFE_SUPPORT) at *any* time point during their listing. Otherwise, zero circulatory points are assigned. A patient is assigned one encephalopathy point if their encephalopathy (CANHX_ENCEPH) is graded to be 1 at the time in question. Otherwise, zero encephalopathy points are assigned. A patient is assigned one INR point if their INR is greater than or equal to 2.5 at the time in question. Otherwise, zero INR points are assigned. Finally, a patient is assigned one bilirubin point if their measured level of serum bilirubin (CANHX_BILI) is greater than or equal to 12 mg/dL at the time in question. Otherwise zero bilirubin points are assigned.

The cumulative ACLF grade is computed by taking the minimum of 3, and the sum of the candidate's renal points, respiratory points, circulatory points, encephalopathy points, INR points, and bilirubin points.

#### Appendix G: Hypothesis Testing

In the main body of the paper, we report several $p$-values associated with statistical tests on our results. This section clarifies our hypothesis testing procedure and presents additional hypothesis test results.

Our general procedure for significance testing is as follows. For each risk model, we perform 1,000 bootstrap resamples (resamples with replacement) from the evaluation set[§] and calculate an evaluation statistic (e.g., 90-day concordance) on each resampled dataset. This gives us 1,000 samples from the distribution of model performance on the resampled evaluation set. These samples are used to perform significance testing (detailed below), and to produce 95% confidence intervals for the bar plots shown in the main body and in the supplement, by selecting the 2.5$^{th}$ and 97.5$^{th}$ percentiles of this distribution to represent the lower and upper bounds of the confidence interval, respectively.

As we are interested in testing noninferiority (e.g., of the collection of DynaMELD models compared to a given baseline), we perform one-trailed $t$-tests. Our analysis makes use of the `statsmodels` Python package's implementation of the one-tailed $t$-test (the `ttest_ind` function)[13].

To formalize this, let $X_{eval}$ denote the evaluation set, and let $X_{eval}^{i,*}$ denote the $i$th bootstrap resampled version of $X_{eval}$. Then, let $C_{model}^{i,eval,*}$ denote an evaluation statistic, $C$ (e.g., 90-day concordance, pooled group concordance), calculated on the resampled dataset $X_{eval}^{i,*}$ for a given $model$ (e.g., DynaMELD;

[§] In this context, we use "evaluation set" to refer to either the *test* set, *pre-COVID test* set, or *post-COVID test* set, as our evaluation procedure is identical across these three sets.

MELDNa). For brevity, we use the notation $\bar{C}_{model}^{eval,*}$ to denote the empirical mean value of $C_{model}^{i,eval,*}$ over 1,000 bootstrap resamples of the $model$ on the dataset $eval$.

$$\bar{C}_{model}^{eval,*} = \frac{1}{1000} \sum_{i=1}^{1000} C_{model}^{i,eval,*}.$$

### Appendix H: SHAP Values

#### Appendix H.1: Clinical Interpretation of SHAP Values

##### *Appendix H.1.1: Model-Agnostic Explanations via SHAP Values*

To interpret the weights of a linear model of the form $y = \beta x$, one can look at the values of the coefficients, $\beta$, to understand the relative significance that the model assigns to each feature when making predictions. By examining the equation of the MELD score, for example, one can deduce that creatinine, bilirubin, and INR are all positively correlated with patient risk, and that in the population on which the MELD was trained, *log*-INR represented a more significant predictive factor than *log*-creatinine, which represented a more significant predictive factor than *log*-bilirubin. Similarly, by examining the equation of the MELDNa, we can additionally deduce that sodium is negatively correlated with patient risk.

The same procedure cannot be directly applied to neural network models. The sequence of matrix multiplications and nonlinear function transformations that comprise the forward pass of the network mean that there is no single network weight representing the significance of a given covariate to the prediction. Moreover, in a sufficiently deep neural network, covariates can interact in arbitrary ways to yield the predicted outcome.

SHAP values represent a procedure to perform feature attribution in nonlinear neural networks. For each data point $i = 1, \dots, n$ and each covariate $j = 1, \dots, m$, the SHAP procedure assigns a real valued number, $\zeta_{ij}$, called a "SHAP value". The SHAP value can be thought of as a surrogate to the linear regression coefficient *for a particular data point*: a positive SHAP value indicates that the value of that data point led the model to yield a larger predicted output. Similarly, a negative SHAP value indicates the opposite: the value of that data point led the model to yield a smaller predicted output. Like linear regression coefficients, SHAP values are an additive means of feature attribution – the difference between the population expected prediction, and the prediction made on a particular data point, can be represented by a sum of SHAP values. However, unlike linear regression coefficients, SHAP values vary across data points, because they represent the marginal influence of each feature in coalition with the other feature values for each given data point.

SHAP values arose from the literature on cooperative game theory[14]; only recently has the concept been applied to feature attribution in machine learning models[15]. At a high level, the procedure works as follows: for each possible combination of features (called a "coalition"), we can examine the marginal contribution of each feature to a given coalition – that is, the average change in prediction when the given feature is added into an existing coalition of features. The SHAP value is a weighted sum of these marginal contributions. As a technical note, the computation of exact SHAP values is computationally expensive: a model must be re-trained once for each coalition, of which there are $2^m$ (one coalition for including / excluding each feature). The `shap` Python package implements a computationally efficient means of computing "SHAP values" (SHAP Additive exPlanations), approximations to SHAP values for neural network models. Due to the number of covariates leveraged in the development of DynaMELD, it is these SHAP values that we present in our analysis.

*Appendix H.1.2: Interpreting a Waterfall Plot of SHAP Values*

In this section, we provide a more detailed interpretation of the "waterfall plot" that we showcased in Figure 4 of the main manuscript. These figures visualize the SHAP values corresponding to a single data point. The plot begins by highlighting the population expected score; equivalently, this can be thought of as the prediction of the model over the trivial coalition (the coalition with no members), which is equal to the expected value of the labels in the data used to calibrate the SHAP procedure. Then, the SHAP values associated with each feature are sequentially added to the expected score, in order from lowest to highest absolute value. The resulting sum is equal to the model's prediction on the data point in question.

Two takeaways are evident from the below plot. First, the relative significance of covariates can change over the course of a patient's trajectory. As we mentioned in the main body, this patient's diagnosis and age represented the third and fourth most significant predictive feature 43 days after listing; by 79 days after listing, these had declined in significance relative to the patient's serum bilirubin and serum albumin. This highlights how – unlike a linear model in which the significance of each feature is fixed over each data point – a neural network-based approach like DynaMELD captures a more nuanced, dynamic picture of the factors driving patient risk.

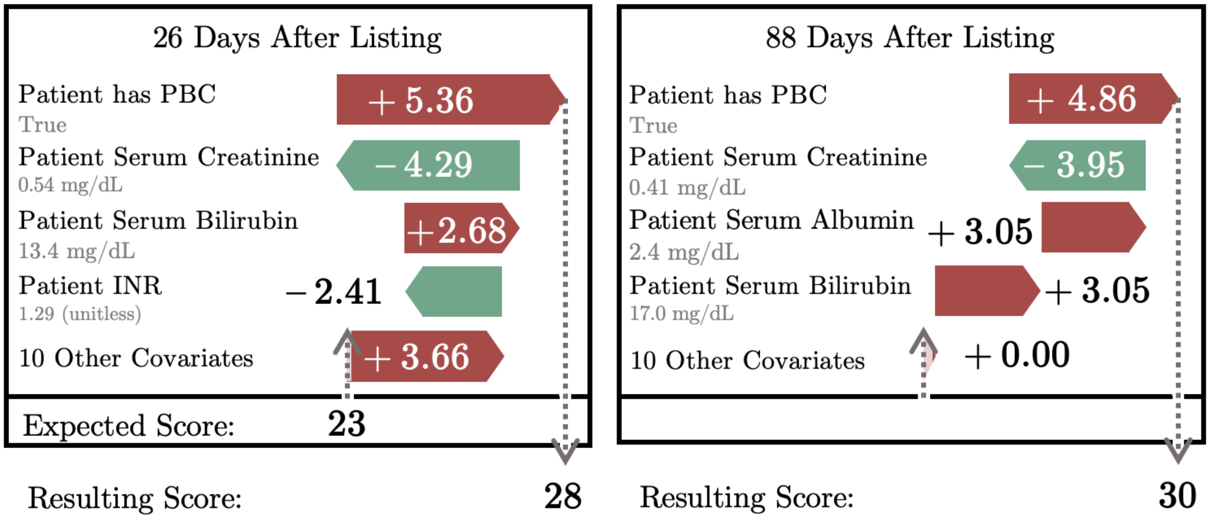

**Figure S.8:** Reproduction of two of the waterfall plots from Figure 4 of the main manuscript, highlighting how SHAP values allow for an interpretation of the DynaMELD model that is individualized to each patient and each observation.
